# Longitudinally Tracking Personal Physiomes for Precision Management of Childhood Epilepsy

**DOI:** 10.1101/2022.11.21.22282474

**Authors:** Peifang Jiang, Feng Gao, Sixing Liu, Sai Zhang, Xicheng Zhang, Zhezhi Xia, Weiqin Zhang, Tiejia Jiang, Jason L. Zhu, Zhaolei Zhang, Qiang Shu, Michael Snyder, Jingjing Li

## Abstract

Our current understanding of human physiology and activities is largely derived from sparse and discrete individual clinical measurements. To achieve precise, proactive, and effective health management of an individual, longitudinal, and dense tracking of personal physiomes and activities is required, which is only feasible by utilizing wearable biosensors. As a pilot study, we implemented a cloud computing infrastructure to integrate wearable sensors, mobile computing, digital signal processing, and machine learning to improve early detection of seizure onsets in children. We recruited 99 children diagnosed with epilepsy and longitudinally tracked them at single-second resolution using a wearable wristband, and prospectively acquired more than one billion data points. This unique dataset offered us an opportunity to quantify physiological dynamics (e.g., heart rate, stress response) across age groups and to identify physiological irregularities upon epilepsy onset. The high-dimensional personal physiome and activity profiles displayed a clustering pattern anchored by patient age groups. These signatory patterns included strong age and sex-specific effects on varying circadian rhythms and stress responses across major childhood developmental stages. For each patient, we further compared the physiological and activity profiles associated with seizure onsets with the personal baseline and developed a machine learning framework to accurately capture these onset moments. The performance of this framework was further replicated in another independent patient cohort. We next referenced our predictions with the electroencephalogram (EEG) signals on selected patients and demonstrated that our approach could detect subtle seizures not recognized by humans and could detect seizures prior to clinical onset. Our work demonstrated the feasibility of a real-time mobile infrastructure in a clinical setting, which has the potential to be valuable in caring for epileptic patients. Extension of such a system has the potential to be leveraged as a health management device or longitudinal phenotyping tool in clinical cohort studies.

**Author Summary:** Epilepsy is the most common childhood neurological condition, affecting 0.5-1% of children worldwide. Affected individuals often have unpredictable seizure events, which, if not timely monitored or treated, can have debilitating or life-threatening consequences. We have developed an early alert system, which is based on wearable devices (e.g., wristband) connected to an adjacent cell phone via Bluetooth. The wearable devices have multiple sensors to collect physiological measurements including heart rate, body movement, and skin responses. These real-time measurements are transmitted via the cell phone to a remote cloud-based computing infrastructure and are compared to the individual’s baseline data. If an abnormal event such as seizure is detected, a message is then pushed to alert the caregiver. In a pilot study tracking 99 epileptic children, we demonstrated that our system was able to detect the onset of seizure events at a high accuracy, often before being noticed by caregivers. Our work demonstrated the feasibility of a real-time mobile infrastructure in a clinical setting, which is valuable in caring for epileptic patients. Extension of such a system has the potential to be leveraged as a health management device or precision phenotyping tool in clinical studies.

## 1. Introduction

### 1.1 Overview

Our understanding of personal physiologies is largely derived from discrete and sparse measurements in standard clinical settings; however, these measurement (e.g., heart rate, blood pressure) are continuous in nature and vary substantially among individuals or potentially change through ages. For example, the baseline body temperature varies from 35.5 to 37.7 Celsius among individuals, therefore using a single threshold (e.g., population average) could potentially complicate clinical decision-making [1]. Similarly, blood glucose level also fluctuates significantly and considerable physiological dynamics underlying dysglycemia among individuals (glucotypes) has been observed in recent studies [2, 3]. Compared with the current standard clinical practice, precision and proactive health management requires dynamically capturing and densely sampling continuous physiological and activity signals, which allows precise definition of health baselines at personal level. As such, illness can be precisely defined, not based on a population average, but by observing significant departures from baseline measurements of the same individual [4]. However, until recently, such a practice was largely infeasible due to technical and implementation obstacles.

These technical challenges can be solved, in part, by re-purposing wearable sensors to achieve longitudinal tracking of physiological signals. Substantial efforts and considerations are required to deploy such a concept in a clinical environment, as outlined below. (1) In contrast to earlier studies on healthy individuals where sensor data were manually exported every a few days for aggregated retrospective analysis [4], deploying a longitudinal tracking system for clinical purpose requires rapid data transmission and synchronization in real time between the front-end sensors for data acquisition and back-end computing infrastructure for rapid decision making. Only systems with rapid information exchange capabilities can achieve personalized diagnosis and intervention in real time. (2) Different from monitoring healthy individuals, clinical deployments usually target specific patient groups, which requires substantial domain expertise to tailor the health management system to meet specific clinical requirements. (3) Clinical deployments also impose higher standards on the monitoring sensors, requiring high sampling-frequency devices to avoid any loss of clinically relevant signals such as brief heartbeat pauses. These technical challenges had limited the implementation of remote and real-time monitoring only to specific situations such as intensive care units (ICU) for life-threatening conditions. In this study, we aimed to transform the standard practice by leveraging cost-effective wearable sensors, building real-time synchronized cloud and mobile computing infrastructures, and developing machine learning platforms.

### 1.2 Predicting seizure onsets on epileptic children

As a proof-of-principle study, we targeted childhood epilepsy, due to the immediately available clinical resources, strong clinical demands and actionability. Epilepsy is the most common neurological condition in childhood, affecting 0.5-1% of children worldwide [5]. It is also a condition commonly comorbid with other neurodevelopmental disorders, including autism and attention deficit hyperactivity disorder (ADHD) [6]. The onset and consequences of epileptic seizures are often unpredictable, causing unexpected injuries and impaired quality of life. Therefore, capturing early signs of epileptic seizure onsets is in strong demand in clinical practice and is particularly critical for infants and toddlers with immature and vulnerable central nervous systems. More importantly, in contrast to the obvious clinical presentation of the generalized tonic-clonic seizures, subtle seizures alone account for approximately half of the seizure events among neonates, which are clinically inconspicuous and difficult to be immediately recognized by caregivers [7]. For epileptic patients in general, many seizure events only last a few seconds or do not present typical forms, e.g., increased motor activity and perspiration and the presence of obvious seizure. These subtle and brief events, adversely affecting brain development in children, cannot be easily captured in the current standard clinical care.

Our long-term goal is to improve clinical practice by building a computing infrastructure for longitudinal <u>o</u>bservation <u>o</u>f <u>p</u>hysiomes (LOOP) for the purpose of data-driven precision health management. We envision that, once developed, and deployed, LOOP can integrate real-time cloud computing, artificial intelligence (AI)-based live clinical decision support, as well as engagement of physicians, patient families and data scientists. In this proof-of-principle study on childhood epilepsy, we re-purposed wearable devices that consisted of multiple sensors capable of sampling physiological and activity signals at a single-second resolution. This system is synchronized with cloud-based back-end infrastructure for real-time communication; machine learning algorithm is trained to detect seizure onsets and caregivers are notified in real time by a front-end mobile app.

### 1.3 Similar studies in early seizure detection

In a clinical setting, standard EEG (electroencephalogram) devices can reliably detect seizure events, but they are usually impractical for longitudinal monitoring. Efforts had been made to record continuous EEG signals by implanting intracranial electrodes [8]; however, the invasive nature of such techniques has limited their usability. Several recent studies also explored using electrocardiogram (ECG) to predict epilepsy; however, the test cohorts usually had fewer than 50 individuals [9-12]. In addition, the higher cost and care of standard ECG devices have limited their potential for wide deployment and longitudinal monitoring in non-clinical settings. Several studies have explored using portable ECG/EEG devices in predicting seizure onsets [10, 13]; however, these studies were often retrospective, and only targeted a very small number of human subjects and could not achieve real-time surveillance and on-time alert.

There were also earlier studies which explored using wearable sensors in seizure detection. The association between GSR (galvanic skin response) and epilepsy-associated cortical excitation was discussed as early as in 2004 [14]; however, its clinical utility for health management was only established on a small set of patients [15, 16]. In a study published in 2017, Onorati and colleagues designed a customized wristband equipped with non-invasive galvanic skin response (GSR) and accelerometer (ACC) sensors and tested the device on a cohort of 22 adult and paediatric subjects diagnosed with epilepsy [17]. Despite reported good predictive performance, this study had the limitation of small cohort size and small number (55) of seizure events captured. It is also not clear how the results can be readily extrapolated to children. Follow-up studies by the same group of researchers expanded the study cohort to 69 patients, totalling 2,311 hours of monitoring time and 452 reported seizure events [18]. Machine learning algorithms were later explored to achieve the best predictive performance [19]. This study had a cohort size similar to ours and also used multiple types of sensors. Despite of the similarities, our work described here is still unique as it deploys a cloud-based system capable of data collection, transmission, and analysis, all in real time. Our proof-of-principle study also used re-purposed wristband (Microsoft Band), which are more accessible than custom-designed wristbands.

## 2. Materials and Methods

### 2.1 Patient recruitment

The study cohort was recruited at Zhejiang University Children Hospital (ZJUCH) Division of Psychiatry between Jan and July 2018. The study was approved by the ZJUCH review board, and guardians of the children were provided written informed consent. These children were all previously diagnosed with epilepsy, and they were admitted to the hospital upon requested by their parents or guardians due to recent seizure episodes. The Inclusion and Exclusion criteria are listed below.

#### Inclusion criteria

Children aged from 0∼18 with focal epilepsy, generalized epilepsy and epileptic encephalopathy were included. Epilepsy was diagnosed by experienced neurology physicians from Children’s Hospital of Zhejiang University, School of Medicine, according to 2017 International League Against Epilepsy (ILAE) classification of the epilepsies [20]. These criteria include: (i) at least two unprovoked (or reflex) seizures occurring >24 h apart, (ii) one unprovoked (or reflex) seizure and a probability of further seizures similar to the general recurrence risk (at least 60%) after two unprovoked seizures, occurring over the next 10 years, (iii) diagnosis of an epilepsy syndrome.

#### Exclusion criteria

(i) Non-epileptic seizures accompanying alterations in consciousness, sensation, motor function, and mentality were excluded, such as syncope, hysterics, transient ischemic attack, hypoglycemia, hypocalcaemia, somnambulism, psychotic disorders, and extrapyramidal diseases, etc. (ii) Participants withdrew due to various reasons during the study were considered excluded.

We also excluded cases when the child strongly resisted the wristband or had other severe comorbidities. The final study cohort consisted of a total of 99 children, which were further randomly divided into discovery (66) and replication (33) cohorts, respectively. Each patient was admitted for v-EEG monitoring at the hospital. Each patient wore the Microsoft Band wristband at all times including during sleep; the wristband was not taken off during physical activities. During most of the time, the patients’ activities were not restricted while at the same time they were encouraged to rest. The patient characteristics are shown in **Table 1;** the detailed characteristics are shown in **S1 Table**. More information on recruitment guidelines is included in **S1 Text**.

**Table 1.** Overview of 66 patients in the discovery cohort

|  | Mean | Min | Q1 | Median | Q3 | Max |
| --- | --- | --- | --- | --- | --- | --- |
| Age (month) | 37.3 | 1 | 6 | 20.5 | 57 | 161 |
| Total duration of time being monitored (in hours) | 84.8 | 1 | 45.25 | 69 | 100.5 | 568 |
| Average duration of time in seizure per day (in minutes) | 6.2 | 0 | 0 | 0.6 | 5.7 | 137.1 |

**Overview of 33 patients in the validation cohort**
|  | Mean | Min | Q1 | Median | Q3 | Max |
| --- | --- | --- | --- | --- | --- | --- |
| Age (month) | 40 | 3 | 4.25 | 10.5 | 72.7 | 156 |
| Total duration of time being monitored (in hours) | 87.7 | 15 | 52.7 | 71 | 109.5 | 208 |
| Average duration of time in seizure per day (in minutes) | 8.7 | 0 | 0.3 | 4.5 | 7.6 | 78 |

### 2.2 Seizure identification

Each child had been previously diagnosed of being epileptic prior to this study. While they were in the hospital, they were monitored around the clock by caregivers or nurses. Due to lack of EEG equipment, for most of the time, a patient was not subjected to EEG measurement; instead, the seizure episodes and other unusual events were observed by the caregivers and recorded by using the app on the smartphone.

### 2.3 Mobile devices and data collection

We elected to use Microsoft Band wristband in this study, which contains 11 types of integrated sensors including an optical photoplethysmogram (PPG) sensor for heart rate (HR), a three-axis accelerometer (ACC) for activity tracking, a galvanic skin response (GSR) sensor for stress detection, as well as a gyrometer (GYR, gesture detection), a GPS, a microphone, an ambient light sensor, a UV sensor, a skin temperature (ST) sensor, a capacitive sensor, and a barometer. Due to practical consideration of battery run-time, we only activated four types of sensors, ACC, HR, GSR, and GYR in this project. The extended run time allowed us to conduct dense sampling and longitudinal profiling. The analyses presented here can be readily extended to signals from any sensor types.

Each wristband is connected via Bluetooth to a nearby smartphone, and the measurements are transmitted to the smartphone at the real time. This smartphone has a pre-installed dedicated app, which allows the caregiver to record it when the child is having a seizure or other unusual events. This smartphone is also connected to a remote central cloud data server via Wi-Fi or cellular network in real time. The recorded data is securely stored on the remote server for storage and analysis. The app has an intuitive user interface, which allows the user to access and display historical data, either recent data stored locally on the cell phone or remotely on the cloud server. In addition to the smartphone placed near the patient, the patient’s real-time and historical data can also be accessed securely through the cloud server by any remote smart phone. The app and cloud server have security features to prevent hacking and data breach. The private data has been anonymized to remove identifiable personal information.

To achieve real-time data acquisition, we used the accompanied SDK/API toolkits and developed an Android app to enable live communications between the sensors and a mobile phone (**Fig 1**). The acquired HR, GSR, ACC and GYR data points had a resolution at 1Hz, 0.2Hz, 8Hz and 8Hz, respectively. Notably, compared with many other alternative consumer-grade devices, our platform had the highest resolution, providing a unique opportunity to capture epileptic events at high resolution, even for the very brief single-second events (i.e., 1Hz for heart rate).

**Fig 1.**
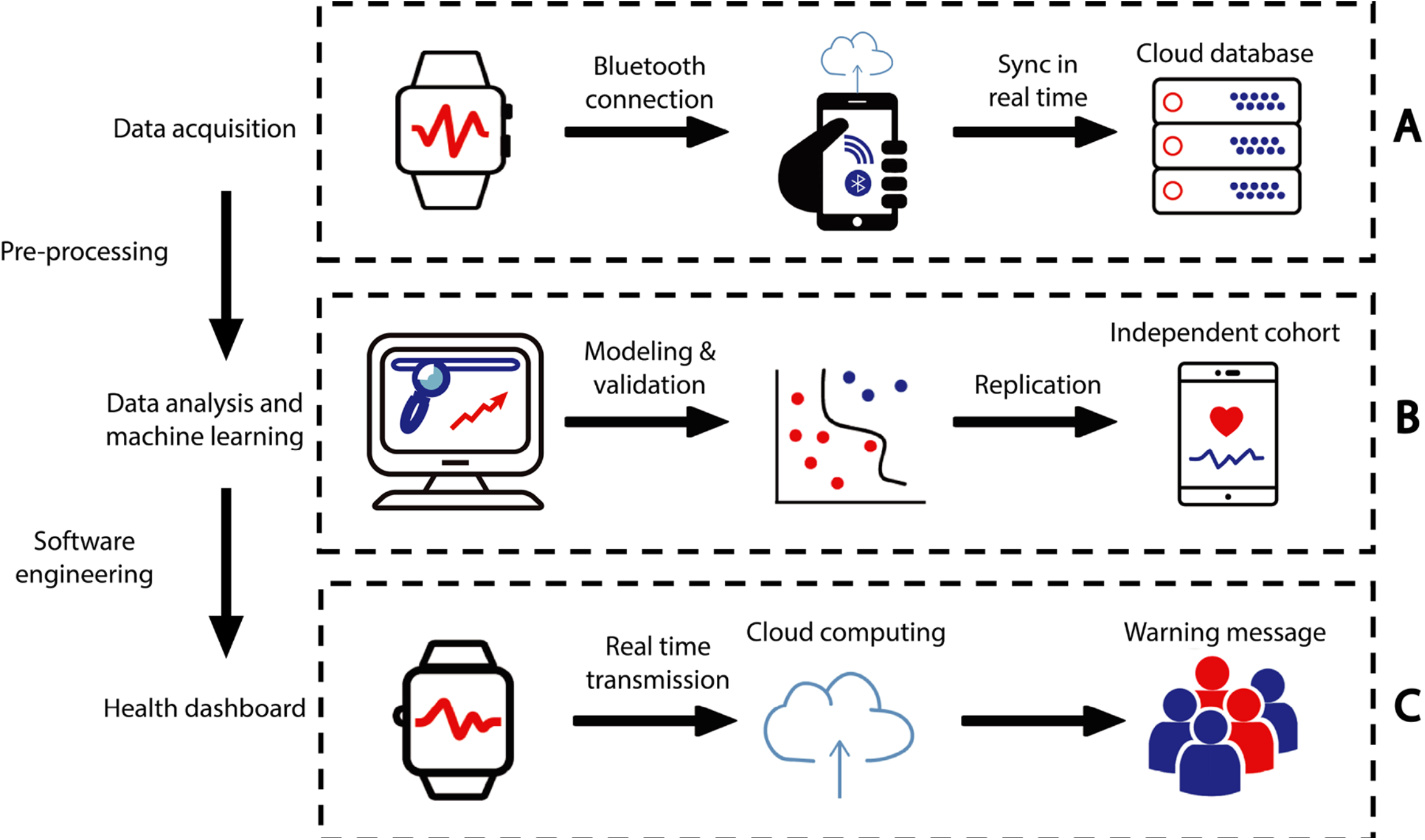
Overview of the LOOP system. The Microsoft Band wristband device is connected via Bluetooth to a nearby smartphone, and the sensor data is transmitted to the smartphone at the real time. The smartphone has a pre-installed dedicated app, which allows the caregiver to record it when the child is having a seizure or other unusual events. This smartphone is also connected to a remote central data server in the cloud via Wi-Fi or cellular network; the recorded data is stored on the remote server for storage and analysis. The smartphone app has an intuitive user interface, allowing the user to access and display historical data, either recent data stored locally on the cell phone or remotely on the cloud server.

During the study, each patient always wore the wristband including during sleep; the wristband was not taken off during physical activities. We discovered that it was more convenient and secure for the children to wear the wristband on their ankles since it was less intrusive to their activities and the device was easily secured. In an earlier similar study, wristbands were also worn on ankles instead of on wrists [18]. During most of the time, the patients’ activities were not restricted while at the same time they were encouraged to rest. In real time, each wristband transmitted the recorded measurements to a connected smartphone, and the smartphone transmitted the data to remote central cloud server for storage and analysis. While in the hospital, the children were monitored around the clock by caregivers or nurses. For most of the time, a patient was not subjected to EEG (Electroencephalogram) measurement due to lack of EEG devices; instead, the seizure episodes and other unusual events were observed by the caregivers and recorded by using an app on the smartphone. We exclusively used Android devices in this study. Patients registered their names at the hospital, which subsequently assigned them a unique user ID and a unique Android device associated with the ID. The Android phones were dedicated for this study, no other apps were installed on the phone and no personal information can be recovered from the device.

### 2.4 Head-to-head comparison between wristband and medical devices on Heart Rates (HR)

To benchmark the sensor data quality, we designed a head-to-head comparison between our heart rate (HR) reads and the measurements from an arm-worn medical grade device (OMRON HEM-6230). We chose one human subject (male, age 20-25, and in good health) for this comparison. We tested each device on this subject under both resting and active states for 30 minutes each. In addition, a trained observer recorded every read change from the arm-worn OMRON HEM-6230 device.

**S1A Fig** compares the HR data acquired from the smart wristband (open blue circles) and from the medical device (red asterisks) in resting state, while **S1B Fig** compares the data in the active state. The measurements acquired by the smart wristband were smoothed by the hardware built-in algorithm; the medical device acquired HR data in every 30 seconds. We next calculated the absolute linearity of these two data series by using the following equation; the measurements from the medical device were deemed as gold standard.

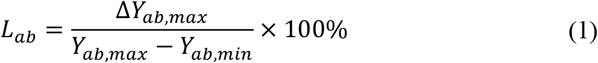

**S1C** and **S1D Fig** show the calculated absolute linearity results in the resting state and in the active state, respectively. The red line is the theoretical linearity measured by the medical device, the cyan dotted line was the theoretical linearity measured by the smart wristband, while the blue dash line is the best-fit line between these two data series. The calculated error rate was 0.4% and 1.2% for the resting and the active state, respectively. Notably, the medical device can only record HR at minute resolution and therefore was unable to capture fast and dynamic HR changes.

### 2.5 Data analysis by PAWO (*Platform for Analyzing Wearable Output*)

We designed a PAWO pipeline (***Platform for Analyzing Wearable Output***) to organize and process sensor signals acquired from the wearable devices (**S2 Fig)**. The input data were longitudinal signals captured by 4 types of wearable sensors at different resolution: a 3-axis accelerometer (ACC, 8Hz), a 3-axis gyrometer (GYR, 8Hz), an optical heart rate meter (HR, 1Hz), and a skin electric response detector (GSR or galvanic skin response, 0.2 Hz). PAWO ensures efficient and robust data transmissions between the wristband and the smartphone, which is capable of handling large volume of data from diverse sensor types. We made the following modifications to the accompanied SDK/API toolkits. (1) We encoded sampling time as 32-bit Unix timestamp accuracy (achieving the resolution to millisecond), providing sufficient resolution for time alignment across events and among data from different sensor types. (2) We packaged sensor type identifiers with the time stamp as well as the acquired sensor readouts into individual data streams, which are subsequently transmitted to the connected mobile phones. (3) We also made the system automatically check the wearing/idle status of the devices, and this signal is separately transmitted to the connected mobile phones. (4) These signals are packaged and sorted automatically at the backend cloud sever.

The PAWO pipeline consists of the following sequential functional modules: Pre-processing, Segmentation, Feature Extractions, and Classification (**S2 Fig**). These individual modules are described in detail below.

#### 2.5.1 Pre-processing module

The pre-processing module was designed for signal quality identification (SQI), denoising, signal smoothing, and adjustment for personal physiological base lines. In the Pre-processing step, these data were cleaned and aligned to the same time axis. The detailed procedures are described below.

##### Signal Quality Identification (SQI)

This module identifies the wearing status of the wristband and removes signals when a device was not active, e.g., during battery charging. This ensures the collection of valid and meaningful data in our study. SQI also filters missing data from the acquired sensor signals. Specifically, we used sliding window method to identify time segments with high rate of missing data. The sliding windows were 60 second in length and non-overlapping. The system removes time segments where more than 60% of the data were missing.

##### Signal Denoising

We adopted discrete wavelet transformation to denoise non-stationary signals such as HR and ACC in this study. The process was divided into the following three steps. (1) Wavelet decomposition of the acquired signals. We implemented wavelet transformation and derived the high frequency coefficients associated with input signals that contained noise. (2) We then thresholded the high frequency coefficients. (3) Wavelet re-construction. We reconstructed the input signal by integrating the thresholded high-frequency coefficients and the originally derived low-frequency coefficients to achieve signal denoising. In our practice, we elected to use DB6 as the wavelet basis function, which has been widely used for processing physiological signals such as heart sound and ECG. We chose to decompose input signals at the level of five when analyzing ACC and GYR signals, where input signals could be decomposed into one low-frequency component and five high-frequency components. Because ACC and GYR had sampling frequencies of 8Hz, the frequency bandwidths of the first to the fifth high-frequency signals were set as D1:2hz ∼ 4Hz, D2:1hz ∼ 2Hz, D3: 0.5Hz ∼ 1Hz, D4: 0.25Hz ∼ 0.5Hz and D5: 0.1Hz ∼ 0.25Hz. The approximate low-frequency bandwidth was 0 ∼ 0.1Hz. The frequency ranges of the acceleration and angular velocity components is 0.3 ∼ 3.5Hz for analyzing epilepsy activities (Poh et al 2012). Based on previous empirical estimate, the frequency ranges from the first to fourth detail layers are expected to be in the range of 0.25Hz ∼ 4Hz, which should contain most of the energy of the input signal. Therefore, we directly set the fifth layer coefficient zero.

The HR signals were first converted into R-R interval (RRI) signal. The frequency distribution of RRI was generally concentrated in the ranges of ultra-low frequency (< 0.04hz), low frequency (0.04-0.15hz) and high-frequency bands (0.15-0.4hz). The bandwidth of high frequency signal components was set as D1: 0.25hz ∼ 0.5Hz, D2: 0.125hz ∼ 0.25hz, D3: 0.06hz ∼ 0.125hz, D3: 0.03hz ∼ 0.06hz. The bandwidth of the low frequency approximation layer was 0 ∼ 0.03Hz. The frequency range of layer 1 to layer 4 was 0.03hz ∼ 0.5Hz., which mostly covered the analysis of HRV from low frequencies to high frequencies.

When thresholding high-frequency components, we selected the hard threshold operator as the threshold function:

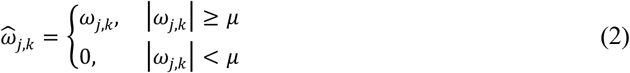

The hard threshold can overcome computing challenges such as singular points in input signals. For seizure detection, singular point signals might contain clinically relevant information. The commonly used VisuShrink method was used to select the threshold [21].

For signal reconstruction, we adopted the Mallat algorithm to decompose input signals, and designed low-pass and high-pass filters to derive the approximate low-frequency and high-frequency coefficients[22]. After thresholding the high-frequency components, we performed step-wise signal reconstruction as follows:

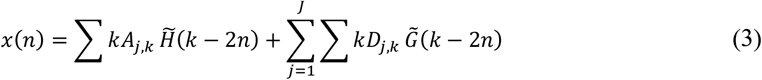

Where 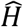. and 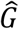 are low-pass filter and high-pass filter respectively, constituting the synthesis filter.

##### Signal Smoothing

We used moving average to smooth signals and removed extreme data points falling the upper and bottom five percentiles across all the signal spectra.

##### Adjusting Personal Physiological Baselines

To account for personal effects on detecting seizure events, we considered personal baseline as the medians of physiological signals from each person and calculated the residue values by subtracting the medians from the input signals.

#### 2.5.2 Segmentation module

In the Segmentation step, we applied standard sliding window techniques to condense and smoothen these longitudinal data [23], i.e., the mean values of the signals were calculated for a window of chosen size. Using statistical analysis and machine learning to detect seizure onsets is essentially a pattern recognition problem for input signals in the time domain. Analyzing multi-channel signals from different sensor types requires unification of their respective time scale, leveraging the time stamps in our UNIX system. We aligned the signals using sliding windows based on the following resolution: (i) For quality check of input signals, the length of each sliding window was one minute, and there was no overlap between any two windows. (ii) For both motion detection and HR/HRV analysis, the length of each sliding window was every five minutes, allowing 80% overlap between adjacent windows.

#### 2.5.3 Feature Extraction module

In the Feature Extraction step, the raw heart rate data (HR) was further processed into HRV (heart rate variation), which quantifies the variation in time between each heartbeat and is a strong indicator of cardiac control of the autonomic nervous system [20, 24]. The HRV associated parameters were further calculated into a set of features, including R-R intervals and SDNN (standard deviation of the normal-to-normal interval), which quantifies the overall variability of heart rate. Notably, SDNN is considered a “gold standard” for medical stratification of cardiac risks [25]. Based on the seizure events manually recorded by nurses or caregivers, the time windows corresponding to seizure onsets were labeled as positive and the remaining windows were labeled as negative. A total of 120 features were extracted from each positive or negative window, including 10 HRV feature set, 20 HR feature set, 80 ACC/GYR feature set and 10 GSR feature set. A bootstrapping-based ensemble network approach was used to remedy the imbalance between negative and positive groups.

The Feature Extraction module has the following components: general analysis to derive overall distributions of the input signals, motion detection for three-axis ACC signals, and detecting abnormal HR/HRV signals.

##### The general analysis component

Statistical analyses were performed to summarize the overall statistical distribution of signals and variables. These statistic data included maximum, minimum, average, median, standard deviation, Q1 (25% quartile), Q3 (75% quartile), 10% percentile, 90% percentile and peak differences.

##### Motion detection from ACC signals

The magnitude of net acceleration was calculated over all three axes of the accelerometer as the following:

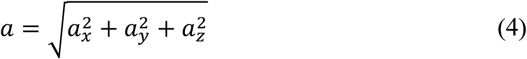

Sliding windows of every 5 minutes with 80% overlap (4 minutes) were used to calculate the standard deviation σ of a given acceleration epoch.

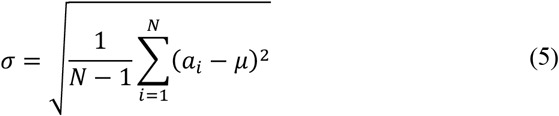

Epochs with σ below 0.1g were considered as resting states and were removed from further analysis. When five-minute sliding window was used allowing 80% overlapping, there were at most 5 identifiers in an epoch. We took the one with more identifier as the epoch identifier.

##### HRV analysis

The HR signals were collected for heart rate viability analysis. Epochs with σ below 0.1 g were automatically discarded from further analysis and treated as non-motor state and hence non-seizure events. Next, R-R interval signals (i.e., distance between consecutive heart beats) were calculated from the valid HR signal through the PAWO pipeline. Finally, R-R interval signal in the 5-minute epochs were used to calculate HRV-associated parameters with sliding windows of 5 minutes with 80% overlap (4 minutes). The HRV-associated parameters used in PAWO included the following: standard deviation of normal R-R intervals (SDNN), root mean squared differences of the standard deviation (RMSSD), adjacent normal R-R interval of difference between the standard deviation (SDSD), the number of pairs of successive NNs that differ by more than 50 ms (NN50), the proportion of NN50 divided by total number of NNs (pNN50) in time domain, very low frequency (VLF, 0.001-0.04Hz), low frequency (LF, 0.04-0.15 Hz), high frequency (HF, 0. 15-0.4 Hz), the ratio of low frequency power and high frequency power (LF/HF), total power (TP) in frequency domain. Normalization was conducted on these HRV-associated parameters against the device build-in smoothing algorithm. When 5 minutes sliding window is used with 80% overlapping, there are at most 5 groups of HRV parameters in an epoch. We take the mean value as the groups of HRV parameters.

#### 2.5.4 Classification module

In the Classification step, the task of seizure detection was posed as a supervised learning task, in which the goal was to classify each 60-second epoch as seizure or non-seizure based on extracted features from physiological signals. The dataset was divided into 10-fold for cross-validation; leave-one-seizure-patient-out cross-validation was used. Area Under Receiver Operating Characteristic curve (AUROC) was used for evaluation. Because of the imbalance of the positive vs negative datasets, Precision-Recall curves were also calculated. The contribution of features was shown in **S3 Fig**. A critical threshold was selected to deem whether an outlier would be considered as an alarm and sent to the mobile end.

### 2.6 Physiome and activity study

The correlation analyses and statistical analyses were conducted on physiological signals to measure physiological changes across childhood developmental stages. Multiple Linear Regression (MLR) analysis was independently conducted on 4 physiological signals (HR, GSR, ACC magnitude and SDNN) by controlling for age, sex, and the time of the day. The pre-processing was performed as follows. First, the time-period in which the human subject had a recorded seizure event were excluded from the analysis. Subsequently, the signals were preprocessed in the PAWO pipelines to denoise; outliners were dropped, and data was normalized by established baselines. In each individual MLR model, the response variables were calculated as the mean of the physiological signals in both daytime and nighttime; and the explanatory variables were age, sex, and the time of the day. The linear model was written as the following:

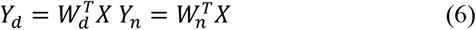

Take HR signal as an example, *Y*_*d*_ ∈ *R*^*R*×1^, *n* = 66 represent the mean HRs of 66 participate in daytime (8 a.m. to 8 p.m.); *Y*_*n*_ ∈ *R*^*n*×1^, *n* = 66 represent the mean HRs in nighttime (9 p.m. to 7.a.m.). Whereas *X* ∈ *R*^*n*×*m*^, *n* = 66, *m* = 3 represented the 3 physiological dimensions (age, sex, the time of the day) of the 66 participants. The above equation could further aggregate into the following:

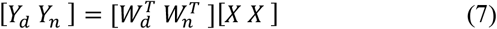

The contribution of age, sex and time were explained via regression coefficients 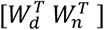; their statistically significances are shown in **Fig 2A**.

**Fig 2.**
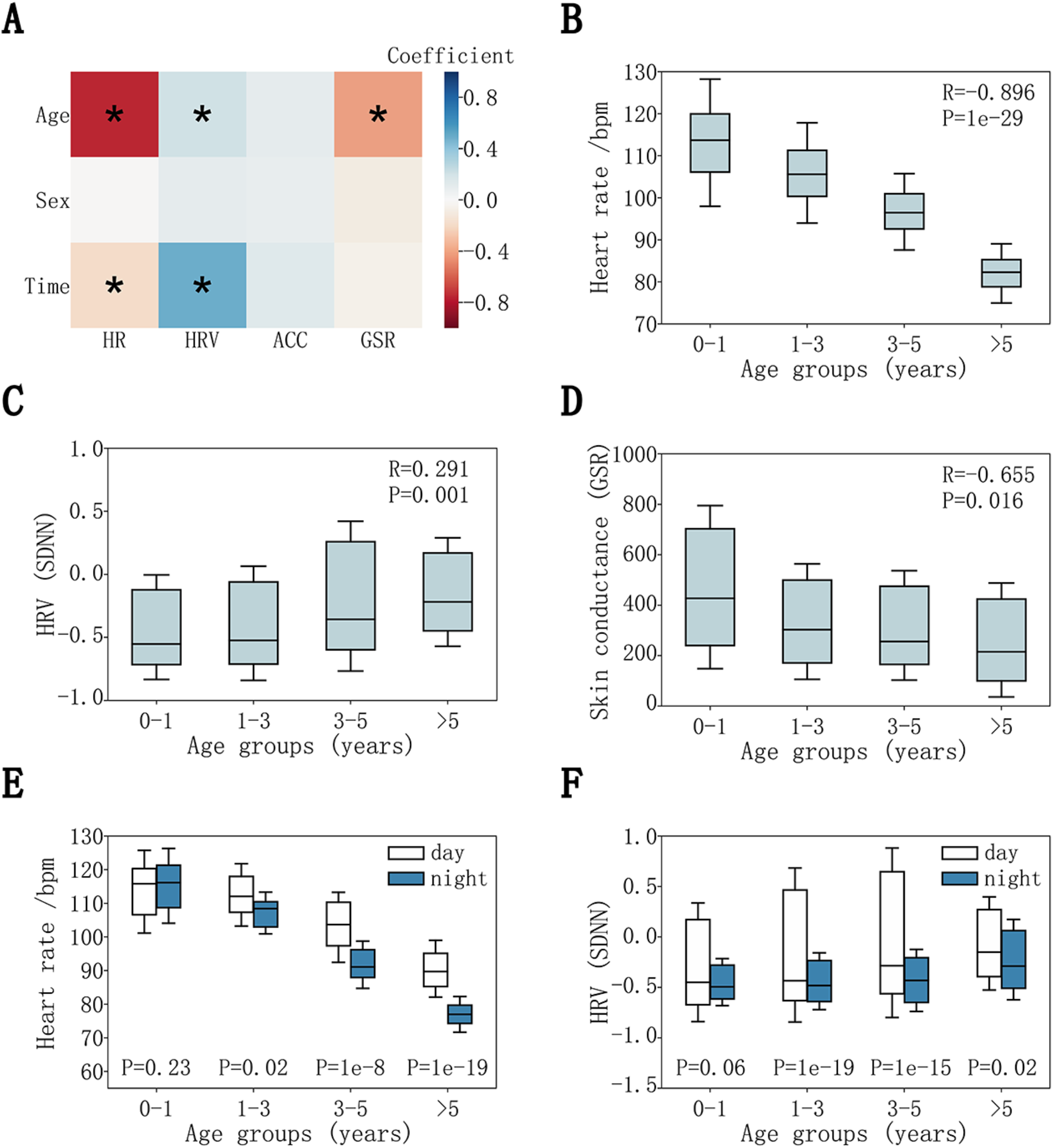
Physiological dynamics associated with childhood development. (**A**) Multiple Linear Regression (MLR) analysis conducted on 4 physiological signals (HR, GSR, ACC magnitude and SDNN variables in HRV analysis) by controlling for age, sex, and the time of the day. (**B**) HR signals associated with childhood development. (**C**) SDNN signals associated with childhood development. (**D**) GSR signals associated with childhood development. (**E**) HR signals associated with childhood development during the time of the day. (**F**) SDNN signals associated with childhood development during the time of the day.

Further statistical analyses were conducted on each physiological signal (HR, GSR, ACC and SDNN) to calculate the distribution characteristics across development stages. Boxplots were used to demonstrate the statistical characteristics, and Wilcoxon rank-sum tests were used to estimate statistical significance. The results were shown in **Fig 2** and **S5 Fig**.

### 2.7 Principal component analysis (PCA) on physiologies during childhood development

PCA is an unsupervised dimensional reduction (DR) technique for mining potential data structure, defined as,

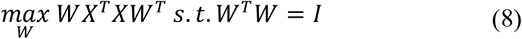

In our study, we implemented PCA to seek clustering pattern of physiology data of the human subjects by mapping from high dimensional space to low visible space, as follows,

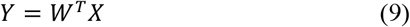

Here, *X* ∈ *R*^*n*×*m*^, *n* = 66, *m* = 70, corresponding to 10 statistical features of 7 variables (HR, SDNN, 3-axis ACC, ACC magnitude, GSR) from 66 participants. *d* was set as 3 in *X* ∈ *R*^*n*×*m*^.

The raw signals from each sensor were processed through PAWO pipeline. SDNN variables and ACC magnitude were also calculated. Statistical analyses were conducted on these 7 groups of variables to acquire 66 × 70 (*n* × *m*) size matrix. Next, before implementing PCA algorithm, zero-mean standardization was applied to normalize features into scale [0 − 1]. Finally, since PCA is an unsupervised method, the algorithm would output 66 three-dimensional 66 × 3 (*n* × *d*) samples in Equation 9 without labeling. The age groups and gender were labeled to highlight the pattern of clustering. The results were shown in **Fig 3**, which indicated clear clustering pattern on age groups while little clustering by gender.

**Fig 3.**
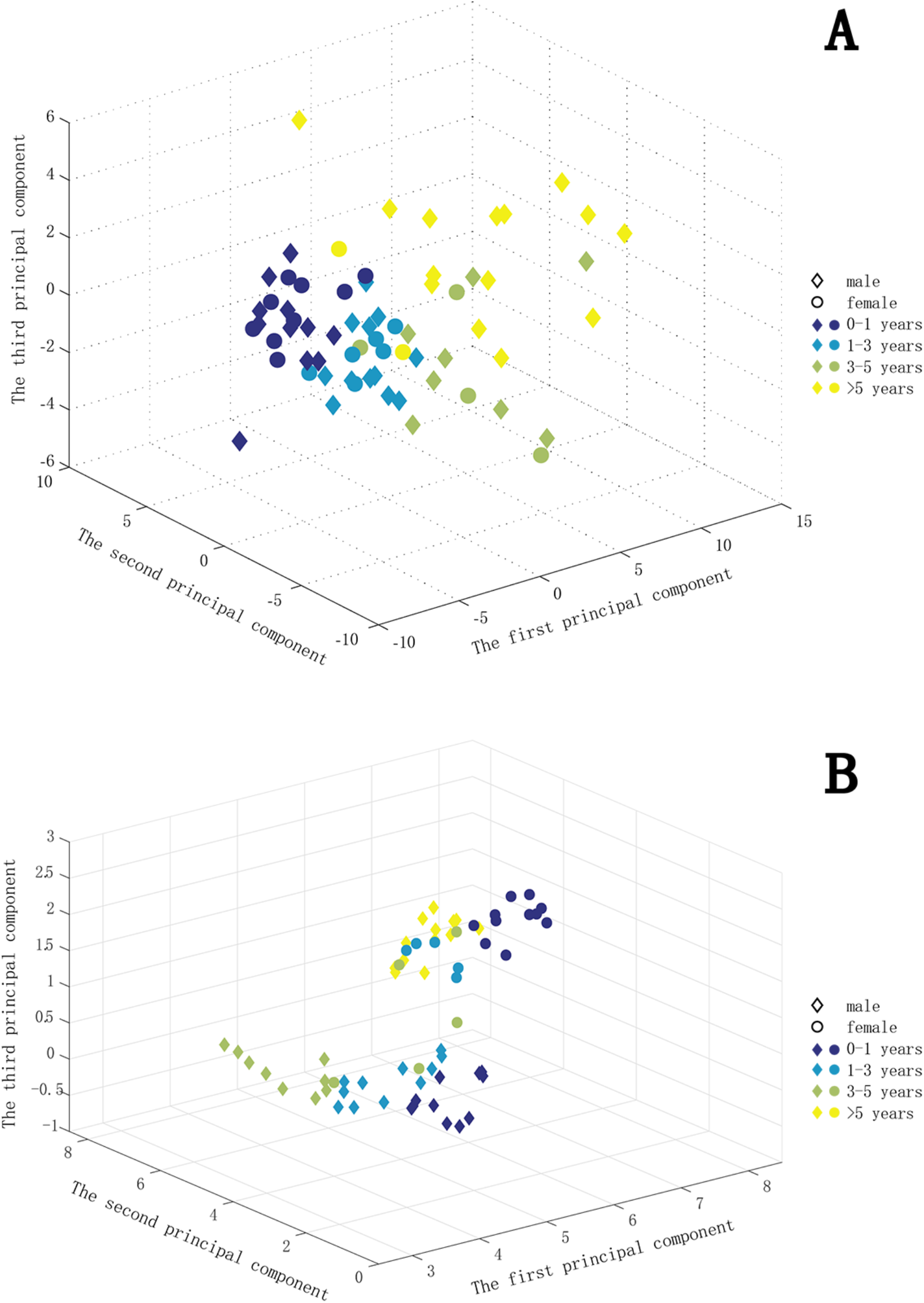
Clustering and visualization of physiological features along the axes of gender and age groups. Each data point represents a vector of a subjects’ HR, ACC, GSR and GYR data. (A) Clustering by Principal Component Analysis (PCA), (B) Clustering results after applying UMAP (Uniform Manifold Approximation and Projection).

### 2.8 Construct prediction model for seizure onset

The overall prediction process was shown in **S2 Fig**. Multiple signals and variables were pre-processed and aligned in PAWO, including 3-axis ACC, 3-axis GYR, HR, GSR, ACC magnitude, GYR magnitude and R-R interval variables. Sliding window technique was applied to condense the data. For each individual, seizure onset times were also added to the window. Statistical procedures and HRV analysis were applied to extract relevant features. A bootstrapping-based ensemble network approach was used to recognize seizure onsets. The problem of seizure detection was posed as a supervised learning task, in which the goal was to classify each 60-second epoch as seizure or non-seizure based on extracted features from EDA and ACM recordings. If any epoch between the start and end of a labeled seizure was correctly classified as a seizure event, the seizure was considered detected (true positive). If multiple epochs within the seizure duration were detected, these were treated as a single true positive. False positives that occurred within 3 minutes from each other were treated as a single false alarm. Details are provided below.

Firstly, sliding window method (60 seconds long) was used to condense the time series into slices. The time window length was set as 60 seconds and overlapping set as 50%, i.e., 30 seconds. The time windows with more than 60% data missing were excluded. Then, according to the seizure events recorded by the nurse or caregivers, the time windows corresponding to seizure onsets were labeled as positive, the remaining windows were labeled as negative. In addition, 60 seconds buffer zones were added to the seizure events; these buffer zones were excluded as negative windows.

Next, features were extracted from each positive or negative window. These features included personal information such as age and sex, time series statistics and HRV variables. The time series parameters included heart rate, R-R interval, 3-axis accelerometer, accelerometer magnitude, 3-axis gyroscope, gyroscope magnitude and galvanic skin response. All the time domain and frequency domain HRV variables were calculated though PAWO. In summary, a total of 120 features from each 60 seconds window was calculated for the subsequent machine learning step to recognize seizure onset.

These features are listed as follows:

1. Heart rate statistics (10): Maximum, Minimum, Mean, Median, Standard deviation, Q1 (25% Quartile), Q3 (75% Quartile), 10 percentile point, 90 percentile point and Peak value difference.
2. R-R Interval statistics (10): Maximum, Minimum, Mean, Median, Standard deviation, Q1 (25% Quartile), Q3 (75% Quartile), 10 percentile point, 90 percentile point and Peak value difference.
3. HRV variables (10): (SDNN), (RMSSD), (SDSD), (NN50), (pNN50) (VLF) (LF), (HF), (LF/HF), (TP)
4. GSR statistics (10): Maximum, Minimum, Mean, Median, Standard deviation, Q1 (25% Quartile), Q3 (75% Quartile), 10 percentile point, 90 percentile point and Peak value difference.
5. Accelerations and the magnitude statistics (40): Maximum, Minimum, Mean, Median, Standard deviation, Q1 (25% Quartile), Q3 (75% Quartile), 10 percentile point, 90 percentile point and Peak value difference.
6. Gyroscope and the magnitude statistics (40): Maximum, Minimum, Mean, Median, Standard deviation, Q1 (25% Quartile), Q3 (75% Quartile), 10 percentile point, 90 percentile point and Peak value difference.

Lastly, a bootstrapping-based embedding network approach, XGBoost [26], was adopted from the sci-learn package [27] and used to overcome the imbalance between negative and positive groups. This model balances the distribution of positive and negative samples by bootstrap sampling and further alleviates the sample bias by embedding independent classifiers. The steps of algorithm are as follows,

1. Construct *n* groups of bootstrap samples (denoted by *S*_*i,i*,=1,2,…*n*_) from the original imbalanced population ***S***.
2. Balance sampling (sample equal number as positive) positive and negative in each group *S*_*i,i*,=1,2,…*n*_. This practice yields a balanced dataset *D*_*i,i*,=1,2,…*n*_
3. Construct stand-alone XGboost classifiers *X*_*i,i*,=1,2,…*n*_ for each dataset *D*_*i,i*,=1,2,…*n*_ as a branch of network.
4. Gather all classifier *X*_*i,i*,=1,2,…*n*_ to generate an ensemble network {*X*_*i*_}_*n*=1,2,…*n*_.

The XGboost classifiers in each branch had equal effect [26]. Therefore, the final prediction score of bootstrapping-based ensemble network was averaged over the prediction scores output by all classifiers, i.e.,

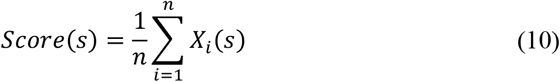

where, *n* is the branch number of ensemble network.

A total of 734,933,434 data points collected from the discovery cohort were used to construct the XGboost model. The dataset was divided into 10-fold for cross-validation; leave-one-seizure-patient-out cross-validation was used. In the whole data set, the patients were divided into ten groups, and the total number of seizure onset was distributed as close as possible among the groups. In each validation process, nine groups were used as the training set, and the remaining one was used as the test set. In the training process, 64 branches were constructed in the bootstrapping-based ensemble network {*X*_*i*_}_*n*=1,2,…,64_. XGboost classifiers in each branch shared the same parameter settings, which were tuned by the random grid-search technique. The configuration parameters were set as the following: number of estimators were set as 400, learning rate was set as 0.1, scale positive weight was set as 1, silent set was set as 0, subsample set was set as 0.9, regulation coefficients were set as 2 and 0, minimum child weight was set as 3, maximum tree depth was set as 8. The testing process was implemented on the trained XGBoost ensemble network.

A critical threshold was selected to deem whether an outlier would be considered as an alarm and sent to the mobile end. This optimal decision threshold was selected at maximal slope at the tangent line of the ROC curve. In this study, the decision threshold was set as 0.5 in the model.

### 2.9 Construction of the cloud-based prediction system leveraging the pre-trained model

An alarm system based on cloud computing was implemented using the trained XGBoost ensemble network with the optimized decision-making threshold. The alarm system possessed the complete training ensemble network {*S*_*i*_}_*i*=1,2,…,64_ from the blind validation procedure. All the samples were subsequently tested on our real-time data. The real-time data steam was continuously transmitted from wearable sensors to the backend cloud. In the cloud sever, PAWO executed the prediction algorithm every thirty second to analyze input signal at a time interval of 60 seconds. With the pre-trained model, we can immediately compute the prediction scores. When prediction scores were beyond our optimized threshold, the system immediately sent out warning messages to caregivers. Fast data transmission, cloud synchronization and automatic prediction has enabled fast decision making almost right after any abnormal physiological signals captured by the machine.

The alarm system was applied on 33 independently recruited children (who were not involved in training the model). A total of 297,651,517 wearable sensor data points from these individuals were screened by our cloud system, and 11 out of the 33 children were recorded at least one seizure event. We compared the systems warning signal against clinical observations as well as against EEG signals whenever possible.

## 3. Results

### 3.1 Overview of the LOOP system

Fig 1 shows the overall implementation of the LOOP system, where sensors are connected via Bluetooth with a mobile phone, which is then synchronized with a cloud sever in real time for data transmission. The raw physiological and activity data were collected, stored in a centralized data warehouse for continuous recording and were subsequently pre-processed in the cloud, where a set of analytic tools were implemented. The clinical decisions based on the machine learning analysis, carried out by the PAWO analysis pipeline (see **Materials and Methods** and **S2 Fig)**, were then pushed to an app on the caregivers’ mobile phone to alert for any seizure onset. The communication among sensors, the cloud severs, the back-end storage and the front-end mobile apps occur in real time at single-second resolution, which can achieve the following functionalities: (1) continuous monitoring of physiological and activity signals from patients, (2) real-time identification of abnormal physiological signals, such as seizure onset, (3) enabling aggregation and storage of historical data to stratify patient cohorts in retrospective studies. Compared with traditional approaches such as patient questionnaires and medical records, which are reactive and discrete, LOOP is proactive and highly individualized.

### 3.2 Physiological dynamics across childhood developmental stages

During the study, we collected a total of 734,933,434 and 297,651,517 sensor data points from the discovery and replication cohorts, respectively, totaling 1.03 billion data points. Given the wide age distribution in our study cohort, this large data resource enabled us to study physiological changes across childhood developmental stages, i.e., at different ages.

We first excluded sensor data acquired between the onset and offset of each seizure event (accounting for 0.2% of the data) and only retained data that were between seizure events. These were data reads associated with normal baseline states. We next regressed physiological and activity datasets with study participants’ age, sex, and the time of the day when signals were recorded. We observed that HR, GSR and HRV signals were significantly associated with study subjects’ age, demonstrating a strong age effect on the measured physiological signals (**Fig 2A**). Interestingly, we observed that HRs were inversely correlated with age (R=-0.896, *P*<1e-29), reflecting the heart developmental process progressing from infants (average HR ∼115 bpm) to children older than 5 years (average ∼80 bpm) (**Fig 2B**). For GSR signals, we observed a negative correlation with age (R=-0.655, *P*<0.016), where the infant group (0-1 years old) displayed substantially elevated GSR (**Fig 2C**). This observation is consistent with elevated reaction to external stimuli or emotional arousal as reflected by skin conductance during infancy, characteristic of the developing sympathetic nervous system during childhood [28]. For HRV, we observed a positive correlation between age and SDNN (standard deviation of the normal-to-normal (NN) interval) (R=0.291, *P*<0.001), where the infants and toddlers displayed the lowest SDNN (**Fig 2D**), also a strong indicator of the developing autonomic nervous system. We also investigated and showed that there were little correlation or dependency among these physiological measurements (**S4 Fig**).

The results of our regression also indicated that HR and HRV signals were correlated with whether the signals were acquired during daytime or nighttime (**Fig 2A**). Except for the infant group of age below 1 year (*P*>0.05, Wilcoxon rank-sum test, **Fig 2E**), we observed significantly increased HR during daytime relative to nighttime (*P*<0.01, Wilcoxon rank-sum test). Similarly, little difference in HRV (represented by the SDNN metric) between day and night was observed among infant or toddler groups, but the elevation of HRV (SDNN) in daytime relative to nighttime was highly significant in all other age groups (**Fig 2F**). We further tested other HRV metrics, including the time domain measures such as RMSSD (square root of the mean squared differences of successive R-R intervals), pNN50 (proportion of NN intervals differing more than 50 ms to the total number of NN intervals), and the frequency domain measures such as low frequency component (LF, 0.04–0.15 Hz), high frequency component (HF, 0.15–0.4 Hz), and low to high frequency ratio (LF/HF ratio). On every measure tested, we consistently observed little difference in HRV between daytime and nighttime among infants or toddlers (**S5 Fig**). Collectively, these observations suggested interesting trends in developing autonomic nervous system and the immature sleep system in young age children. We also tested the ACC and GYR data, but we did not observe significant association between these data types and the study subjects’ attributes or physical activities. The lack of association can be likely explained by the immobile status of the study subjects that were confined to hospital beds.

Next, to obtain an overview of the wearable sensor data of the cohort, we represented each study subject by his or her own longitudinal physiome and activity profile captured throughout the study period (HR, GSR, ACC), and projected the high-dimensional data onto a three-dimensional space using principal component analysis (PCA) or Uniform Manifold Approximation and Projection) (UMAP) (**Fig 3**). The PCA analysis revealed a clustering pattern of the 66 study participants in the discovery cohort, where young, middle aged, and older children formed separate groups, regardless of gender (**Fig 3A**). In contrast, the UMAP clustering further grouped the patients according to male or female (**Fig 3B**).

Notably the infants (age 0-1) were tightly clustered and closer to the toddlers (age 1-3), displaying a clear separation from children of age 3-5, and even further from children of age>5. This observation is consistent with the developmental-stage-specific physiologies during childhood development, which had been previously reported in the literature [29-34]. Overall, these results suggest that a wearable platform such as described here provides an effective and flexible strategy for longitudinal large cohort studies.

### 3.3 Departure of physiological sensor data from baseline during seizure events

The longitudinal dataset allowed us to define individual physiome baselines over a long period of time; thus, any significant deviation from the baselines would be considered as health concerning. We hypothesized that these datasets would allow early detection of seizure onsets. To do so, we need to first annotate all the true seizure events manually in the patients from both the discovery and the validation cohorts; the recorded seizure events in the discovery cohort will be used to train our machine learning algorithm and the recorded seizure events in the validation cohort will be used to benchmark our algorithms. As described in **Materials and Methods**, each admitted patient is monitored 24/7 by a dedicated nurse or caregivers. Upon a seizure onset, the nurse or the caregiver immediately recorded the event by simply tapping a mobile app (**Fig 1**, panel A). A subset of seizure events was also confirmed by EEG measurements; however, given the limited resources, not every patient was subjected to EEG monitoring. **Table 2** shows the duration of all the seizure events recorded in the discovery and replication cohorts.

**Table 2.** Duration of seizure events (averaged over all patients)

| Duration | < 1 minute | 1-2 minutes | 2-3 minutes | 3-4 minutes | 4-5 minutes | >5 minutes |
| --- | --- | --- | --- | --- | --- | --- |
| Percentage | 81.3% | 10.3% | 3.2% | 2.3% | 1.1% | 1.8% |

Before designing and training our machine learning classifier, we first explored whether the physiological data acquired by the wearable devices indeed can distinguish seizure from baselines. We chose one patient as an example (age 6-12 month), who had two seizure attacks in a ∼4 min interval as recorded by the accompanying nurses (**Fig 4**). As shown in **Fig 4A** (upper panel, time intervals TI1 and TI2), the heart rate (HR) data from this individual precisely recapitulated these two seizure events, hallmarked by two major HR peaks during seizure occurrences. We next mapped the time-course signal to the frequency domain using Short-time Fourier Transformation (STFT), which further revealed high-frequency HR dynamics in the two time-intervals coinciding with seizure onsets (**Fig 4A**, middle panel). We next aligned the electroencephalogram (EEG) data along the same time axis, which revealed that the first and second event started at the 120^th^ and 431^st^ second, respectively, each lasting ∼10 seconds (**Fig 4A**, bottom panel). We further examined each seizure event individually and discovered characteristic epileptic signals such as spike discharges, spikes and sharp waves continued with a rhythm in theta band on Channel P4, T4 and T6 for both events (**Fig 4B** for TI1 and **Fig 4C** for TI2). Five abnormal EEG signals were highlighted in TI1, and three in TI2. Given these observations, we concluded that both detected epileptic events were focal epilepsies originated from the right parietal and temporal lobe of the patient. Because focal epilepsies typically exhibit milder symptoms than generalized epilepsies [35], this case study demonstrated the sensitivity and specificity of our wearable tracking system in capturing even milder epileptic events.

**Fig 4.**
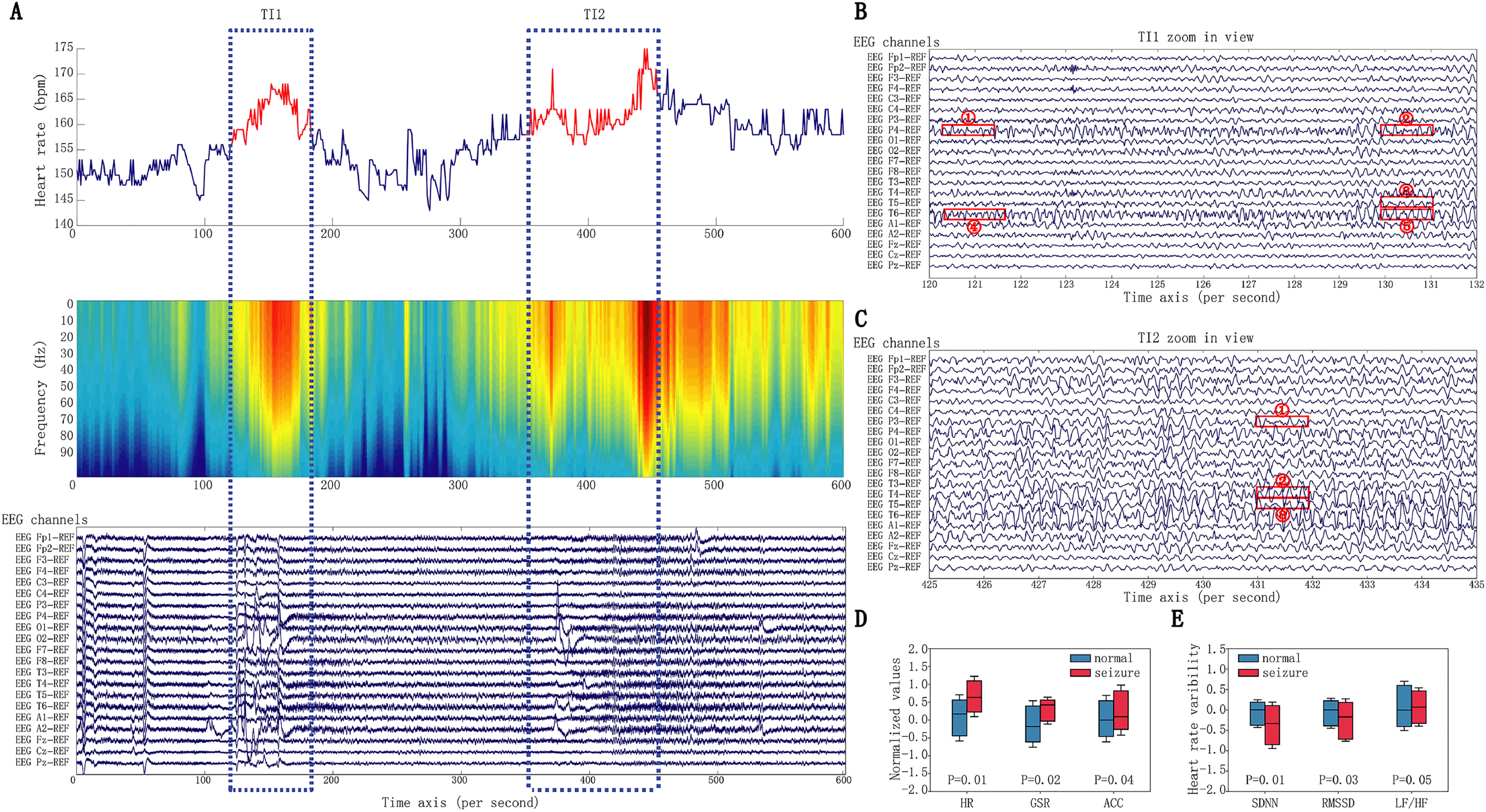
Physiological dynamics associated with seizure events. (**A**). Top panel: Heart rate measurements from a patient in a period of 600 seconds. The measurements during a short time seizure are marked in red, while the signals in the normal periods are marked in blue. Middle panel: The time series measurements were converted by STFT to frequency domain. Bottom panel: EEG signals in the same period are shown in 21 channels. (**B**) Zoomed in view of EEG signal in time interval 1 (TL1). (**C**) Zoomed in view of EEG signal in time interval 2 (TL2). (**D**) The measurement of HR, GSR, ACC during seizure significantly deviated from normal periods. (**E**). The measurement of SDNN, rMSSD, HF/LF during seizure significantly deviated from normal periods.

In addition to heart rate data, we also examined whether individual subjects had abnormal GSR (galvanic skin response) or ACC (axis accelerometer) signals during these confirmed seizure events. For each individual and for each sensor signal type, we subtracted that individual’s median signal level across all the time points; significant departure from the respective personal baselines would be flagged as abnormal. As shown in **Fig 4D**, individuals during seizure onsets indeed tended to have elevated HR (*P*<0.01, Wilcoxon rank-sum test), higher GSR (*P*<0.02) and higher ACC (*P*<0.04). The elevated HR and GSR signals during seizure events were anticipated, and the increase in ACC was likely explained by seizure-associated jerky body movements. We further extended the comparison to the normalized HRV features: SDNN (standard deviations of NN intervals), RMSSD (root mean square of successive differences between normal heartbeats), and LF/HF (Low / High frequency). SDNN (*P*<0.01, Wilcoxon rank-sum test) and RMSSD (*P*<0.03) displayed a significant reduction during seizures relative to normal conditions, whereas LF/HF exhibited a significant increase (*P*<0.05, Wilcoxon rank-sum test) (**Fig 4E**). Because the LF and HF components in HRV reflect sympathetic and parasympathetic nerve activities, respectively, the increased LF/HF thus suggests a shift towards sympathetic dominance, resulting in imbalance of the autonomic nervous system during seizure interictal period [36]. Overall, we concluded that there were significant departures in HR, GSR, GYR, and ACC data from personal baselines during confirmed seizure events, which made it feasible to develop a machine learning method to integrate these data and predict seizure onsets.

### 3.4 Wearable platform and machine learning enable early detection of seizure onsets

Having established significant departures in sensor data from baselines during seizure events, we next sought to build a machine learning framework to predict seizure onsets from these sensor data. For a given individual at a given time point, we extracted its associated HR, GSR, GYR, and ACC signals and calculated their residuals by subtracting respective personal baselines, which served to orthogonalize the recorded signals across different individuals (see **S1 Text** and **S2 Fig**). Briefly, after pre-processing using wavelet packet transform (WPT) and imputation techniques for noise smoothing and filtering, we discretized the high-resolution time-course sensor data into time windows at a step of every 60 seconds. We next calculated the summary statistics of HR, GSR, ACC and GYR data in each time window as well as their Short-time Fourier transformed frequency spectrums. Specifically, for HR data, we calculated their derived HRV measures (e.g., SDNN, R-R intervals) in each time window. Throughout the entire study, 1,290 total seizure events were timed and recorded from the discovery cohort, and we labeled their associated measurements as positive samples.

To overcome the challenge that negative samples (the time windows in which children did not have seizures) outnumbered positive samples, we employed bootstrapping-based balanced sampling technique [37]. The results were evaluated by ten-fold leave-one-seizure-out cross-validation. As noted in the literature, this strategy was a rigorous evaluation using non-overlapping individuals instead of using non-overlapping time intervals from the same individual [17]. We followed the standard practice of using Sensitivity (Sens) and False Alarm Rate (FAR) to assess the performance of our machine learning model [17], where Sens was defined as percentage of recorded seizures that were also identified by the machine learning algorithm, and FAR was defined as the number of false alarms in a 24-hour window. The resulting FAR/Sens pairs corresponding to varying classifier decision thresholds were used to build receiver operating characteristic (ROC) curves in **Fig 5A**. The optimal decision threshold was selected to maximize sensitivity and minimize false alarm rate, i.e., the point close to the top left-hand corner. As shown in **Fig 5A**, our algorithm achieved 89% sensitivity (1,148/1,290 seizures detected) and the average number of false alarms per 24h was 0.54 (125 false alarms in total 5,543 hours). **Fig 5B** shows the ROC curve of the 11 patients in the validation cohort. Because of the imbalance of the positive vs negative datasets, Precision-Recall curves were also calculated (**Fig 5C, 5D**).

**Fig 5.**
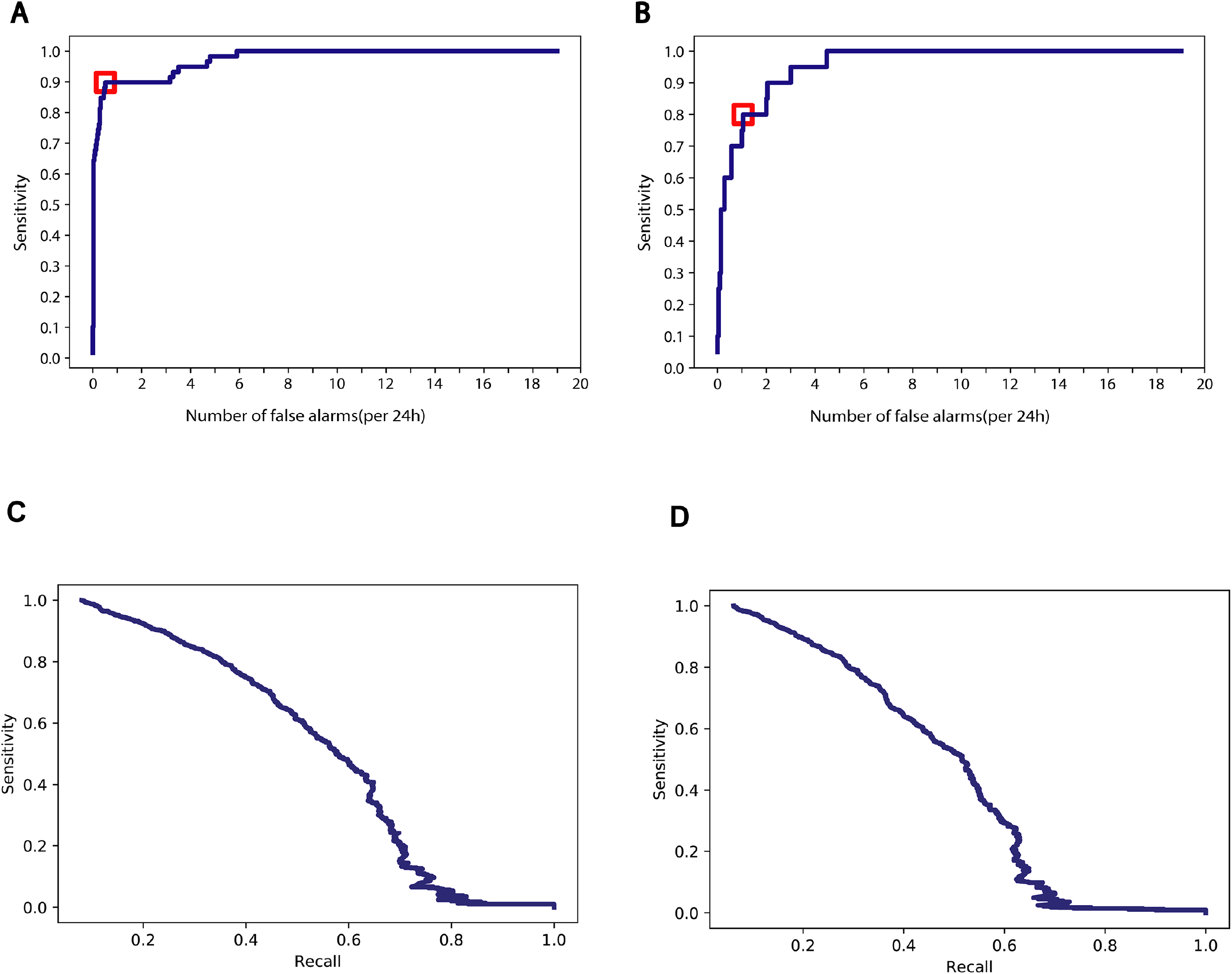
Performance of the machine learning model. (**A**) ROC curve of 10-fold leave-one-seizure-out cross-validation on 66 patients, the threshold is marked in red. (**B**) ROC curve of real-time testing on recorded seizure events of 11 selected subject in the validation cohort, the threshold is marked in red. (**C**) Precision-Recall curves of the prediction accuracy. (**D**) Precision-Recall curves of the validation cohort.

To ascertain the robustness of our prediction, we applied the same prediction algorithm to the replication cohort consisting of 33 children (see **Table 1B)**. Among these 33 children, 35 independent seizure events from 11 children were recorded by observing nurses. With the same model configuration learned from the discovery cohort, we achieved prediction on the replication cohort at 80% sensitivity (28/35 seizures detected) with an average number of false alarms at 1.1 per 24h (10 false alarms in total 264 hours) (**S3A** and **S3B Fig**).

Because the positive seizure events were observed and recorded by nurses or caregivers, we suspected that these were mostly strong and obvious seizure episodes, and subtle seizure events not easily recognized by humans were potentially under-reported. However, if these subtle seizure events triggered physiological irregularities, they would likely be captured by our LOOP system. Along this line, we next examined whether there were seizure events predicted by our model, but not observed by nurses. Indeed, there were many of these instances. One of many examples is shown in **Fig 6A**, where a seizure event was predicted (red bar in the grey box) by our model, but not observed by caregivers. The yellow bars adjacent to the true seizure event represent “false alarms”, which was the result of persistent perturbed physiological data. This patient was on EEG monitoring during the predicted seizure events, so we were able to retrieve EEG measurements aligned with the event axis as shown in **Fig 6B**, where the time interval for the predicted seizure event was shaded in grey. A high-resolution view is presented in **Fig 6C**, which clearly revealed EEG signals characteristic of local epilepsies on Channel P4, T4 and T6. This demonstrated that our approach has the capability to capture events not observed by humans and suggested that the prediction accuracy we reported here was likely an under-estimate.

**Fig 6.**
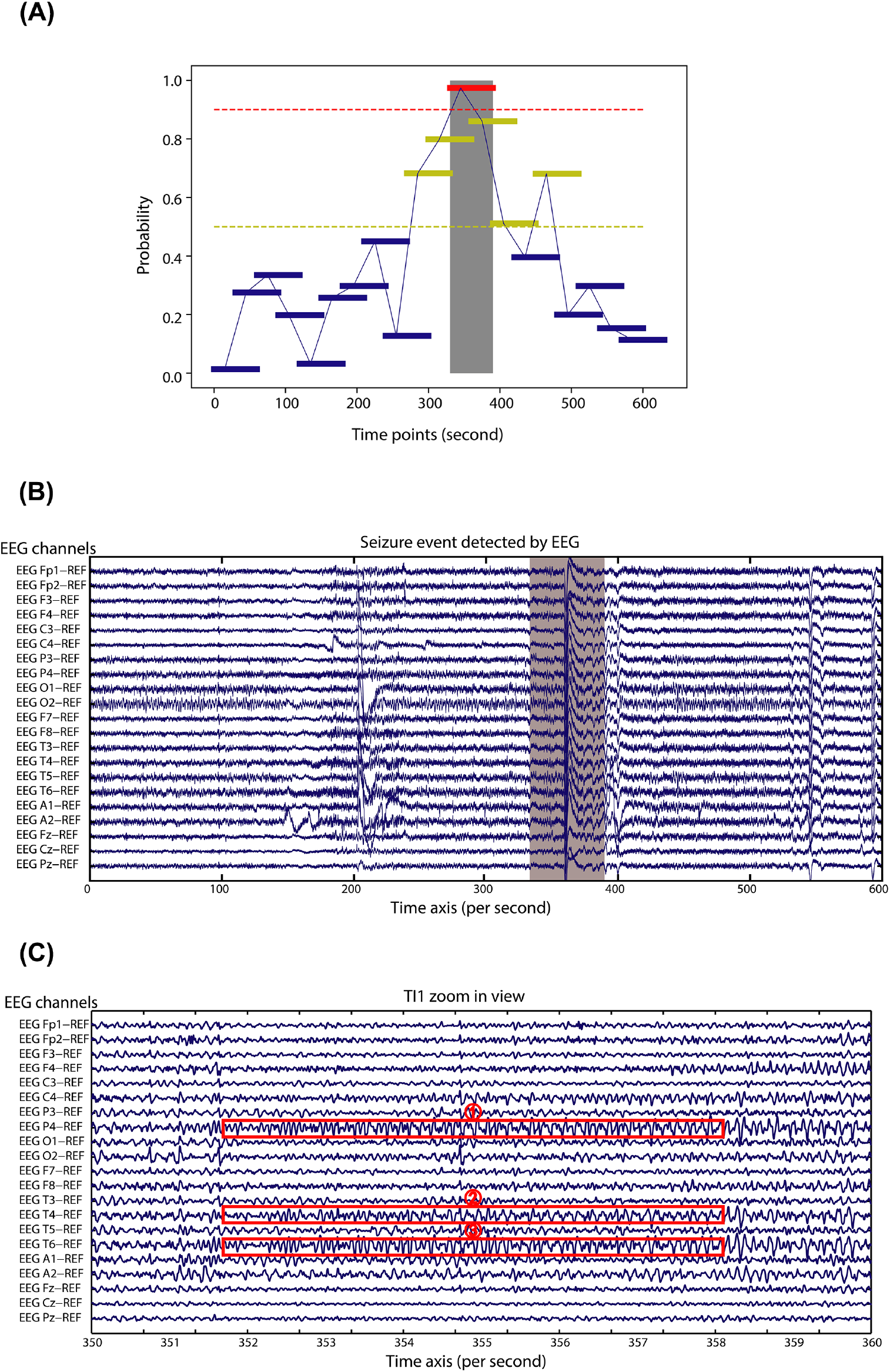
Real-time monitoring during a seizure event. (**A**) Real-time monitoring during a seizure event. The red horizontal bar in the grey box represents a true seizure event, the dark yellow horizontal bars represent “false alarms”, and the blue horizontal bars represent true negative events. (**B**). EEG signals during a seizure event. (**C**). Zoomed in view of EEG signals during the seizure event (TI1).

### 3.5 Building a Personalized Health Dashboard for Management of Epilepsy Conditions

To facilitate potential clinical use of our system, we have built a personalized health dashboard, which enables caregivers to monitor patients’ physiological conditions in real time. The dashboard system can be deployed on computers or on mobile devices. We also implemented our machine learning algorithm on the health dashboard, where real-time predictions are made in the cloud, and communicated to caregivers (**Fig 7**). We have demonstrated its clinical utility in epilepsy in this study; however, this system can be easily adapted to many other clinical scenarios. At the backend, wearable sensor data are stored in a secure data warehouse, which can be leveraged as a precision phenotyping tool for large-scale retrospective analysis on healthy or disease cohorts.

**Fig 7:**
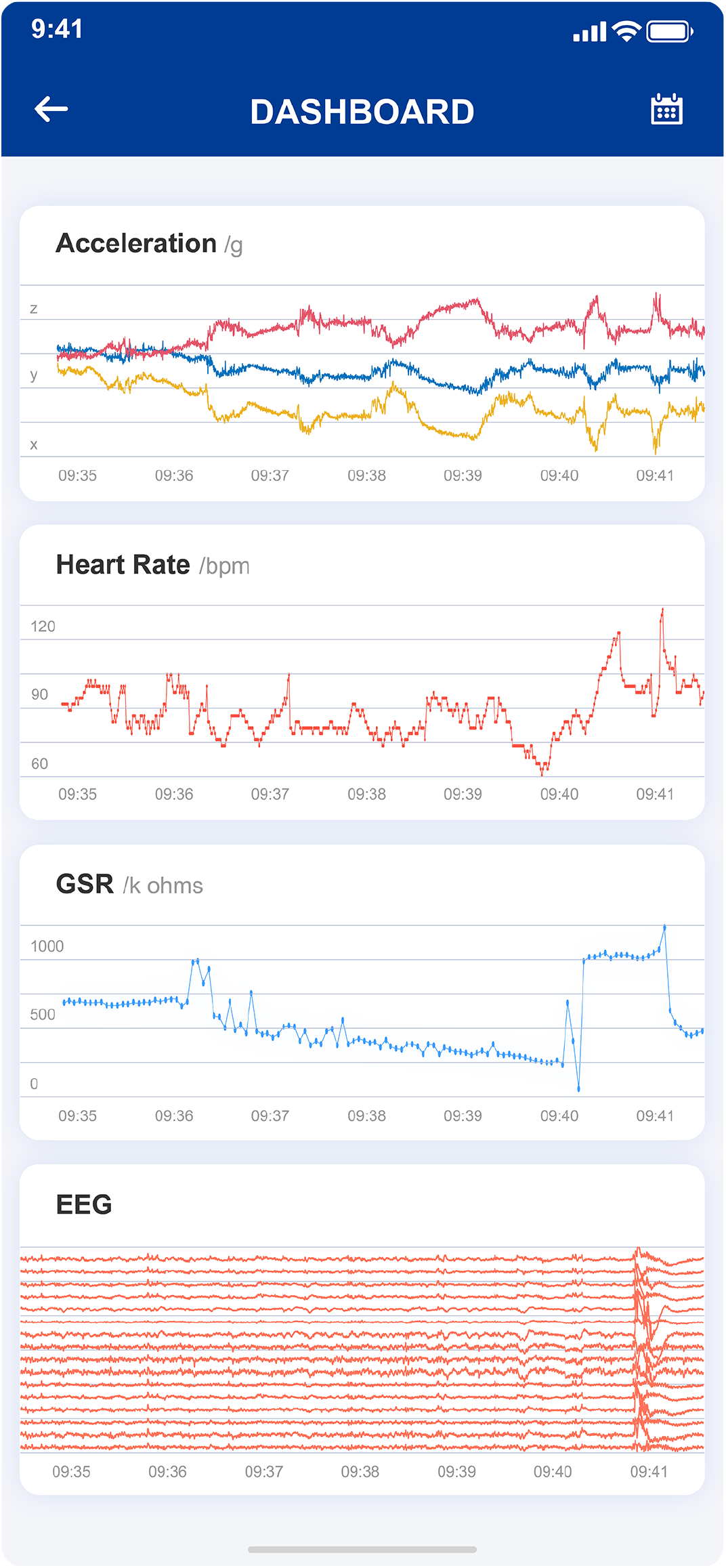
The App interface of LOOP system.

## 4. Discussion

Precision health management aims to improve clinical practice by introducing personalized solutions to predict, prevent and manage human diseases [3]. As such, the first step towards this overarching goal is to enable personalized data acquisition and analytics. Compared with conventional approaches using clinical notes and questionnaires, wearable sensors connected to a cloud computing system represent important technological advancement that enables one to track individuals’ personal activity and physiologic parameters proactively and longitudinally to identify clinically concerning events [4, 38-40]. The large-scale dataset at the personal level brings great opportunities to study clinical traits, and their aggregation will significantly advance public health research.

We have built a cloud computing system, LOOP, which has dynamically acquired more than one billion data points from wearable sensors worn by epileptic children. The LOOP implementation integrates multidisciplinary expertise involving hardware design, software development, cloud computing, digital signal processing and machine learning techniques, as well as the engagement among data scientists, clinical practitioners, and patient families. The work described here demonstrated that, as a proof-of-principle, this system can effectively provide early detection of seizure onsets on epileptic children. To the best of our knowledge, this was one of the largest and dedicated studies applying wearable sensor in childhood epilepsy, albeit there were earlier similar studies on adult epilepsy with smaller cohorts [17-19].

Utilizing our platform, we analyzed physiological dynamics during childhood development, which revealed distinct patterns of HR, GSR and HRV among children, particularly among infants (**Fig 2**). Because GSR is independently controlled by the sympathetic nervous system [41], and HR and HRV are regulated by autonomic nervous system on cardiac rhythm [42], these observations can be explained by the developing autonomic nervous system in children. Interestingly, while we observed that HR and HRV are mostly indistinguishable between daytime and nighttime among infants, the GSR signals at nighttime displayed a consistent and significant reduction relative to the daytime GSR readouts across all age groups. This suggested reduced psychophysiological arousals at nighttime even for infants. Interestingly, in our analysis, we consistently observed increased GSR signals in females relative to males across all age groups, suggesting sex-specific stress responses. Some of our observations, especially those with HR and HRV, are in good agreement with previous studies [35, 43]. Notably, our longitudinal data at single-second resolution provided better granularity in quantifying e physiological differences. Moreover, many of the observations made in this study were not previously reported in the literature, which can only be discovered using high-resolution longitudinal data across a wide age range.

Compared with the common practice of using population average to define health status, our study demonstrated the advantage of using personal physiological baselines to identify clinically concerning events that significantly deviated from such baselines. We demonstrated this concept on childhood epilepsy by analyzing the residual signals departing from personal baselines and developed a machine learning framework to predict seizure onset. The robustness of the framework was further validated in an independent replication cohort. With our longitudinal high-resolution data, we were able to quantify its predictive power in a large cohort. In fact, we observed that it was the HRV and HR-associated parameters that contributed the most to seizure prediction accuracy (**Fig 4D, 4E** and **S3C Fig**). **S3C Fig** shows the top 20 features that contributed the most to the prediction accuracy. The features associated with HRV are shown in dark blue, features associated with HR are shown in light blue, and GSR associated features are show in red. Among the top 20 highest contributing features, eight are HRV features. In addition, the top feature, SDNN, has substantial higher contributions to the prediction accuracy than other features. Intriguingly, we observed a significant reduction of SDNN, RMSSD and an elevation of LF/HF accompanied with seizure onsets, which is consistent with previous studies that were conducted using regular ECG devices [44, 45]. These observations suggested perturbed sympathetic to parasympathetic activities implicated in epilepsy, highlighting the accuracy of our devices. We particularly note that HRV represents HR dynamics, which requires high-resolution data acquisition and real-time data analysis.

Our study has the following limitations and potential room for improvements. It is well known that epilepsy is a complex and heterogeneous neurological disorder which are classified into many subtypes based on clinical manifestation and physiologic diagnosis such as by EEG. Generally, epileptic patients also have comorbidities, which in turn can influence physiological measurements. We did not focus on classifying the study subjects into clinical subtypes; however, we did remove those subjects who had shown extreme comorbidities or had resisted wearing the wristband. We based this decision on two considerations. The first reason was the limited cohort size, which did not provide the luxury of further stratifying the patients into subtypes, despite this study representing the largest childhood epileptic study of this kind. The second reason is that our machine learning classifier was trained on the normal data acquired from the same individual, which minimized confounding factors originated from the heterogeneity among the study subjects. In future studies when larger cohorts become available, it would be very valuable to evaluate the performance of our approach after stratifying the study subjects.

The second limitation is that the confirmed “true positive” seizure events were recorded by accompanying nurses and caregivers, which potentially could introduce its own biases or errors. As we have shown, in many cases, our wearable device could identify seizure events that were missed by humans. This means that the benchmarking metric of our method is probably an underestimate, i.e., many of the “false alarms” were probably real subtle seizure events. EEG (Electroencephalography) is the gold standard in epilepsy diagnosis, unfortunately it was not feasible to conduct EEG on patients in a large cohort 24/7. There were some studies using invasive intracranial EEG devices, which are not practical at the present but may be useful once improved sufficiently[8, 46].

The third limitation is the device itself, which was effective and accurate however was not specifically designed for seizure detection or for children to wear. Other researchers have designed dedicated wearable devices for epilepsy, e.g., Empartica wristband, but these devices are too costly to deploy as consumer product at the moment [17, 19]. New generation of affordable and adjustable wristband are being brought to the market; it would be intriguing to investigate whether these dedicated devices perform better than re-purposed smartphones like what we used in this study.

In summary, we have built a personalized digital health dashboard for dynamically monitoring individual physiome and activity at single-second resolution. This digital health platform, together with machine learning-based clinical decision support system, facilitates the development of personalized prediction and intervention strategies for health management. We note that HR alone is a strong predictor of many clinical conditions, such as cardiovascular, metabolic, and diabetic diseases [47, 48]. Therefore, the developed LOOP system can be adjusted and extended to many other diseases. In addition to health management, since large-scale clinical traits can be acquired using this technology, the system can also serve as precision phenotyping tool, allowing integration with genotyping data.

## Data Availability

All data produced in the present study are available upon reasonable request to the authors.

https://figshare.com/projects/Wearable_Seizure/141719

## Supporting Information

**S1 Text**

Supporting information.

**S1 Table**

Characteristics of the patents tracked in this study.

**S1 Fig.**
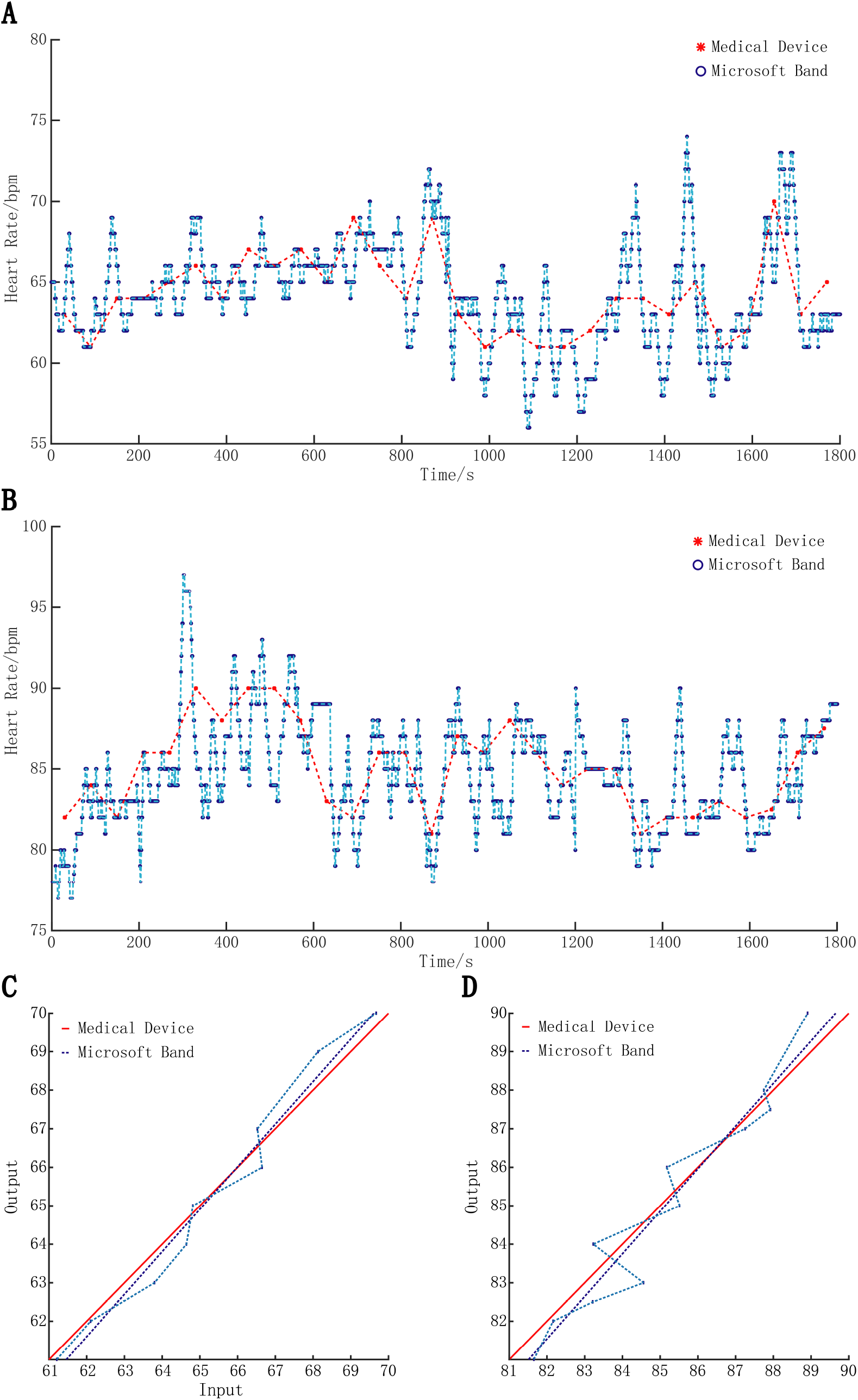
Benchmarking the HR measurements from the smart-band (Microsoft Band 1 and 2) and a medical grade device (OMRON HEM-6230) in the resting state (**A**) and in the active state (**B**). (**C**) The linearity analysis in the resting state, (**D**) The linearity analysis in the active state.

**S2 Fig.**
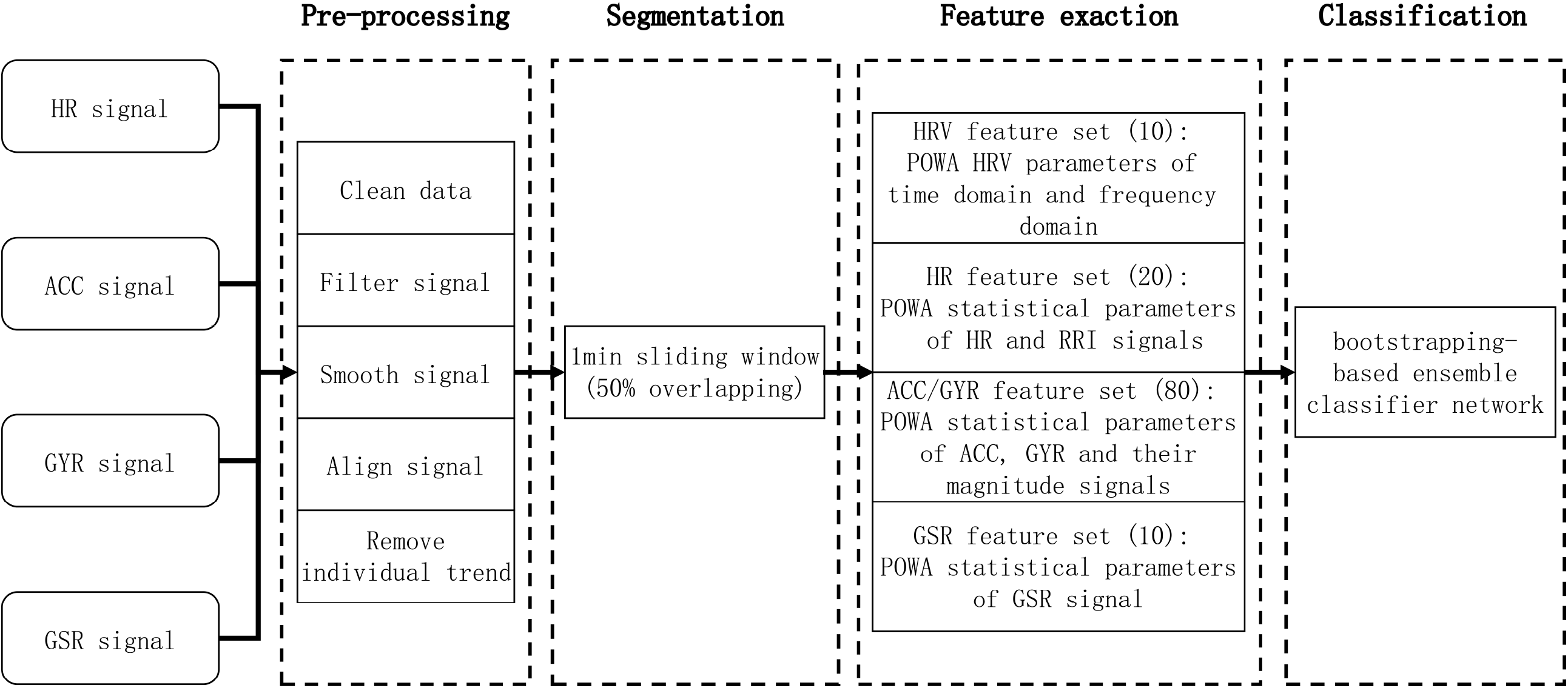
Overview of seizure detection machine learning architecture, PAWO (Platform for Analyzing Wearable Output). The pipeline consists of the following sequential functional modules: Pre-processing, Segmentation, Feature Extractions and Classification.

**S3 Fig.**
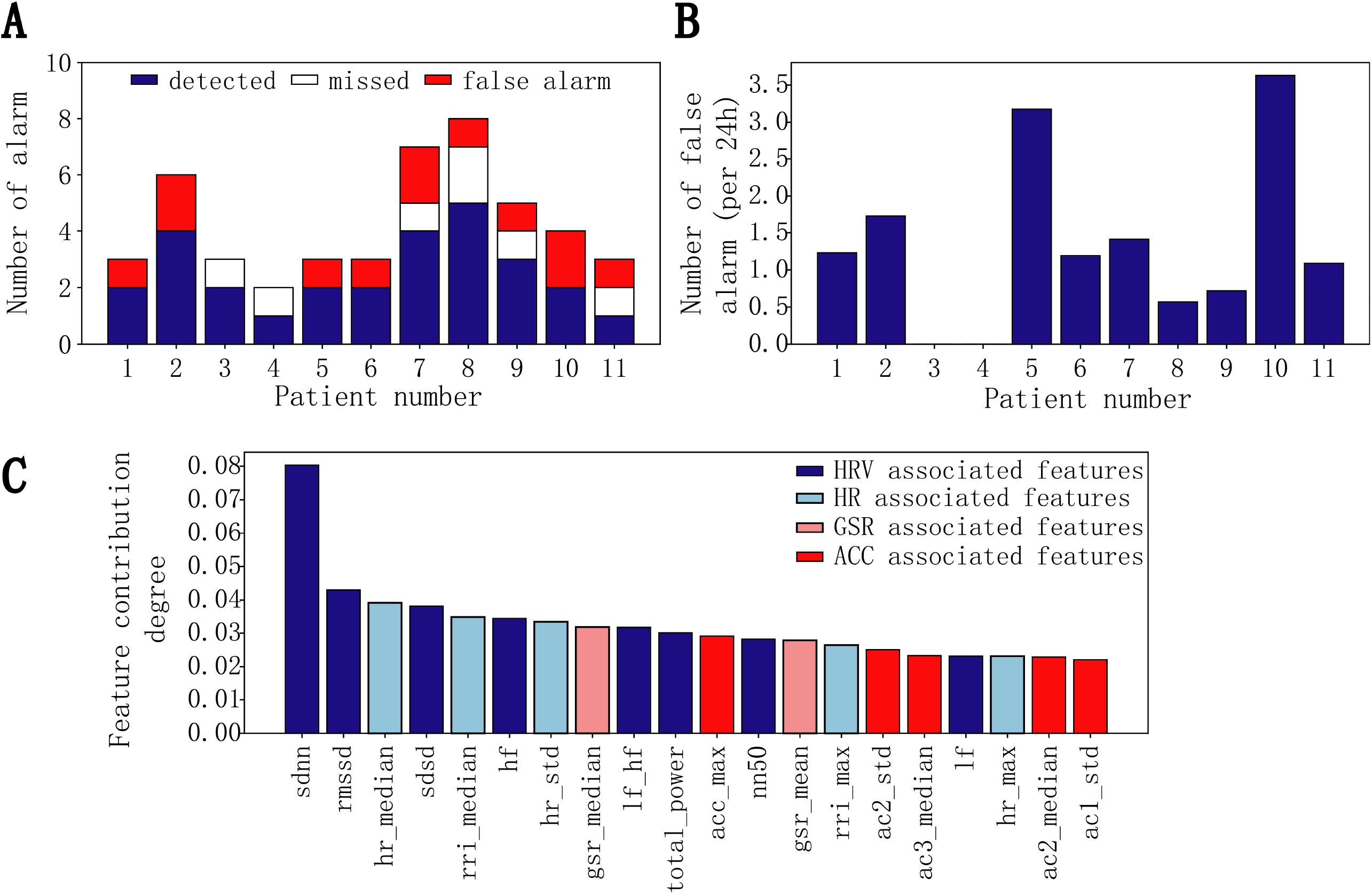
Benchmarking results on the Validation cohort. (**A**) Predicted seizure events from the 11 tested patients. Blue bars represent true positive seizure events as validated by the human observations, white bars represent seizure events recorded by human but missed by predictions, red bars represent false alarms, i.e., predicted seizure events but not observed by humans. (**B**) The number of false alarms per 24 hours on the tested 11 patients. (**C**) Estimated importance of the features in our seizure prediction model, in descending order of top 20, were SDNN, rMSSD, median of HRs, SDSD, median of R-R interval, HF, standard deviation of HRs, median of GSR, LF/HF, TP, maximum of ACC magnitude, NN50, mean of GSR, maximum of R-R interval, standard deviation of ACC in y axis, median of ACC in z axis, LF, maximum of HRs, median of ACC in y axis and standard deviation of ACC in x axis.

**S4 Fig.**
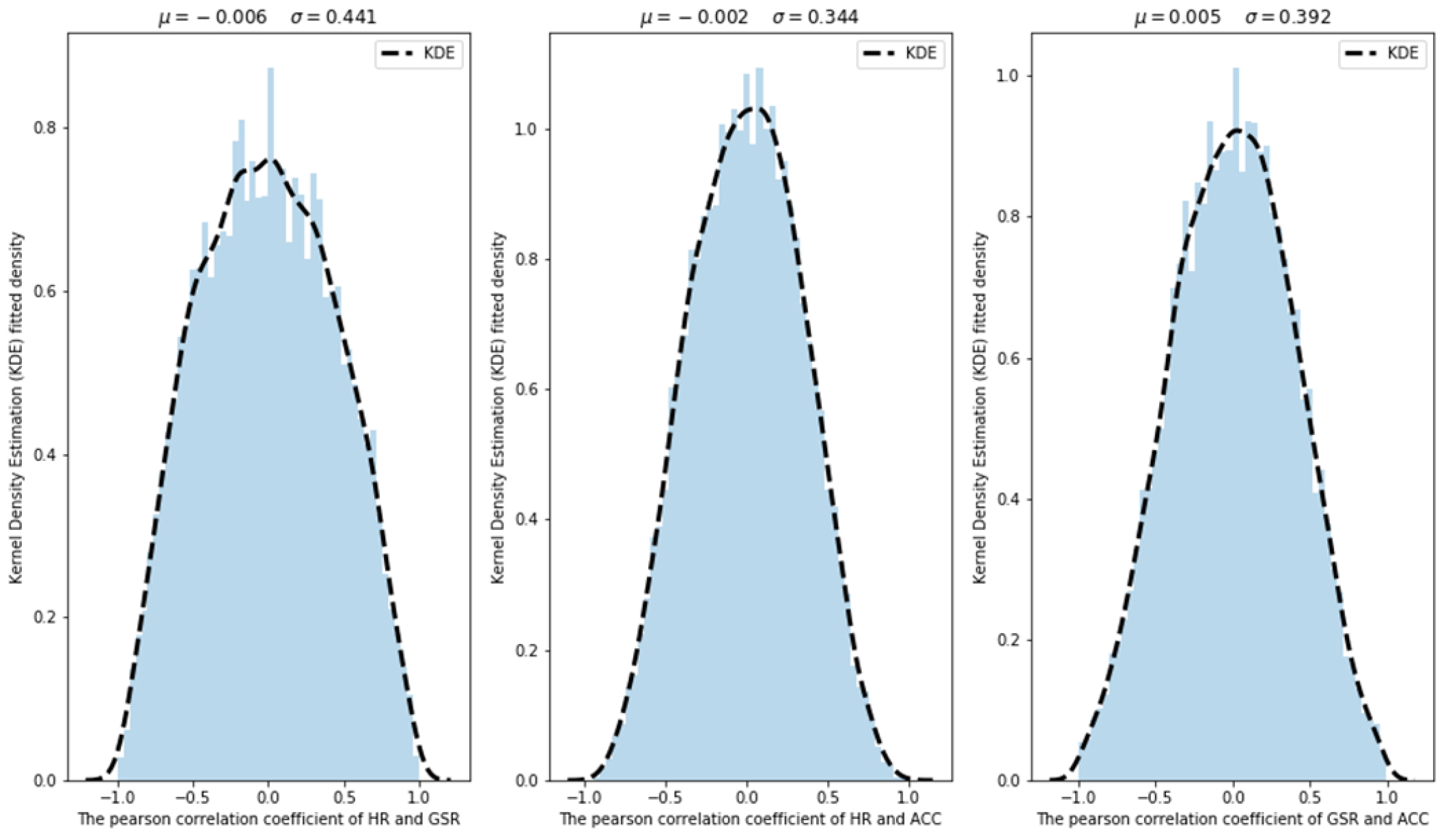
Pearson Correlation Coefficients plots between three types of sensor measurements. The plots show that there is minimal correlation between HR, ACC, GSR.

**S5 Fig:**
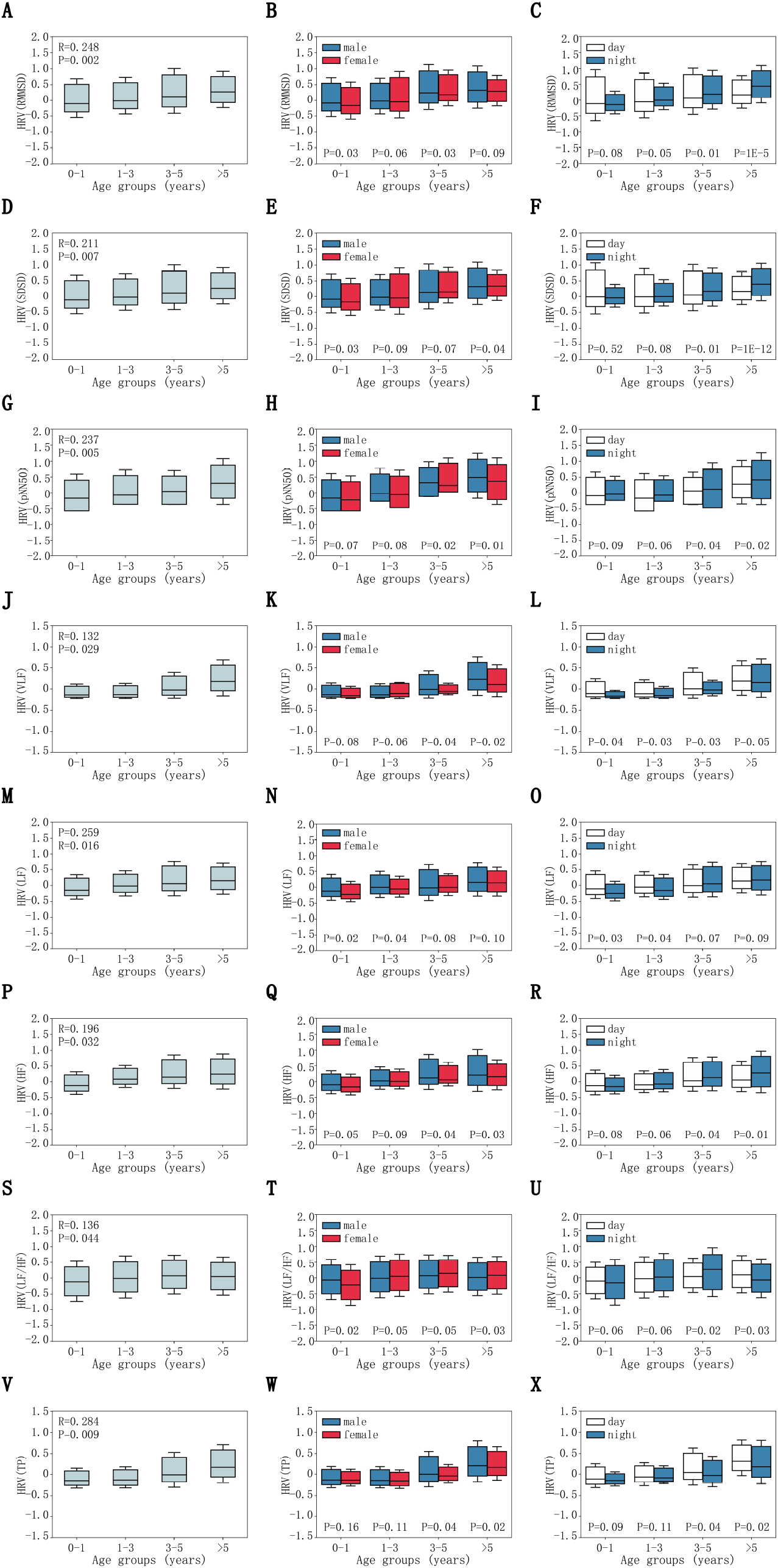
HRV (Heart Rate Variation) dynamics associated with childhood development after controlling for age, sex, and the time of the day. (**A**)-(**I**): time domain HRV associated parameters, (**J**)-(**X**): frequency domain HRV associated parameter. (**A**) rMSSD controlling for age. (**B**) rMSSD controlling for sex. (**C**) rMSSD controlling for the time of the day. (**D**) SDSD controlling for age. (**E**) SDSD controlling for sex. (**F**) SDSD controlling for the time of the day. (**G**) pNN50 controlling for age. (**H**) pNN50 controlling for sex. (**I**) pNN50 controlling for the time of the day. (**J**.) VLF controlling for age. (**K**) VLF controlling for sex. (**L**) VLF controlling for the time of the day. (**M**) LF controlling for age. (**N**) LF controlling for sex. (**O**) LF controlling for the time of the day. (**P**) HF controlling for age. (**Q**) HF controlling for sex. (**R**) HF controlling for the time of the day. (**S**) LF/HF controlling for age. (**T**) LF/HF controlling for sex. (**U**) LF/HF controlling for the time of the day. (**V**) TP controlling for age. (**W**) TP controlling for sex. (**X**) TP controlling for the time of the day.

## Supporting Information

## 1. Patient recruitment guideline and overview

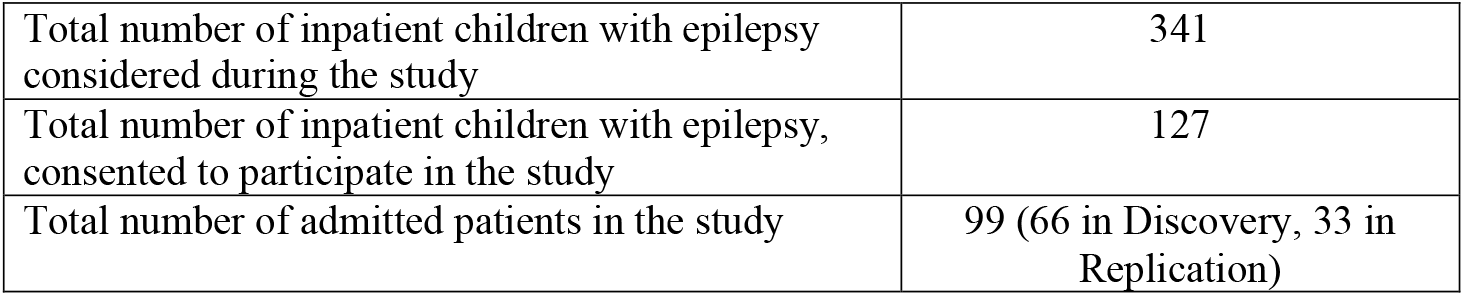

|  |  |
| --- | --- |
| Total number of inpatient children with epilepsy considered during the study | 341 |
| Total number of inpatient children with epilepsy, consented to participate in the study | 127 |
| Total number of admitted patients in the study | 99 (66 in Discovery, 33 in Replication) |

### Inclusion criteria

Children age from 0∼18 with focal epilepsy, generalized epilepsy and epileptic encephalopathy were included; epilepsy was diagnosed by experienced neurology physicians from Children’s Hospital of Zhejiang University, School of Medicine, according to 2017 International League Against Epilepsy (ILAE) classification of the epilepsies (doi: 10.1111/epi.13709) described as follows:

1. At least two unprovoked (or reflex) seizures occurring >24 h apart;
2. One unprovoked (or reflex) seizure and a probability of further seizures similar to the general recurrence risk (at least 60%) after two unprovoked seizures, occurring over the next 10 years
3. Diagnosis of an epilepsy syndrome

### Exclusion criteria

1. Non-epileptic seizures accompanying alterations in consciousness, sensation, motor function, and mentality were excluded, such as syncope, hysterics, transient ischemic attack, hypoglycemia, hypocalcemia, somnambulism, psychotic disorders, and extrapyramidal diseases, etc.
2. Participants withdrew due to various reasons during the study were considered excluded.

The final study cohort consisted of a total of 99 children. They were further divided into discovery (66) and replication (33) cohorts, respectively. The study was approved by the ZJUCH review board, and the parents of the children provided written informed consent. These children were all previously diagnosed with epilepsy, and they were admitted to the hospital upon requested by their parents and guardians due to recent seizure episodes. Each patient was admitted for v-EEG monitoring at the hospital. Each patient wore the Microsoft Band wristband at all times including during sleep; the wristband was not taken off during physical activities. During most of the time, the patients’ activities were not restricted while at the same time they were encouraged to rest.

## 2. Data collection by wristband

Two types of smart-bands were used in this study, Microsoft Band 1 and Band 2, which were equipped with a microphone, 3-axis accelerometer, 3-axis gyroscope (Microsoft Band 2 only), light sensor, thermometer, UV sensor, capacitance sensor, optical heart rate meter, GPS, skin electric response detector (Microsoft Band 2 only) and barometer sensors. Each wristband is connected via Bluetooth to a nearby smartphone, and the measurements are transmitted to the smartphone at the real time. This smartphone has a pre-installed dedicated App, which allows the caregiver to record it when the child is having a seizure or other unusual events. This smartphone is also connected to a remote central cloud data server via Wi-Fi or cellular network in real time. The recorded data is securely stored on the remote server for storage and analysis. The App has an intuitive user interface, which allows the user to access and display historical data, either recent data stored locally on the cell phone or remotely on the cloud server. In addition to the smartphone placed near the patient, the patient’s real time and historical data can also be accessed securely through the cloud server by any remote smart phone. The App and cloud server have security features to prevent hacking and data breach. The private data has been anonymized to remove identifiable personal information.

## 3. Seizure identification

Each child had been previously diagnosed of being epileptic prior to this study. While they were in the hospital, they were monitored around the clock by caregivers or nurses. Due to lack of EEG equipment, for the majority of the time, a patient was not subjected to EEG measurement; instead, the seizure episodes and other unusual events were observed by the caregivers and recorded by using the App on the smartphone.

## 4. Head-to-head comparison between smart-band and medical devices on Heart Rates (HR)

We compared Microsoft Band with an arm-worn medical grade device, OMRON HEM-6230, for its accuracy in measuring heart rate (HR). We chose one human subject (male, age 20-25, and in good health) for this comparison. We tested each device on this subject under both resting and active states for 30 minutes each. In addition, a trained observer recorded every read change from the arm-worn OMRON HEM-6230 device.

**Supplemental Figure S1A** compares the HR data acquired from the smart wristband (open blue circles) and from the medical device (red asterisks) in resting state, while **Figure S1B** compares the data in the active state. The measurements acquired by the smart wristband were smoothed by the hardware built-in algorithm; the medical device acquired HR data in every 30 seconds. We next calculated the absolute linearity of these two data series by using the following equation (**Equation 1**); the measurements from the medical device were deemed as gold standard.

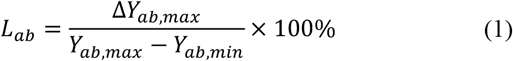

**Supplemental Figure S1C** and **S1D** show the calculated absolute linearity results in the resting state and in the active state, respectively. The red line is the theoretical linearity measured by the medical device, the cyan dotted line was the theoretical linearity measured by the smart wristband, while the blue dash line is the best-fit line between these two data series. The calculated error rate was 0.4% and 1.2% for the resting and the active state, respectively.

## 5. PAWO pipeline

We designed PAWO (***Platform for Analyzing Wearable Output***) pipeline to organize and process sensor signals acquired from the wearable devices, which consisted of four coherent modules: *stream processing, pre-processing, signal analysis and multi-signals time alignment*. The table below outlines key technical details in the platform. The individual modules are described in detail below.

|  | ACC | GYR | HR | GSR |
| --- | --- | --- | --- | --- |
| **Frequency** | 8Hz | 8Hz | 1Hz | 0.1 Hz |
| **Preprocessing** | Yes | Yes | Yes | Yes |
| **Additional analyses** | Motion detection | - | HR/HRV analysis | - |
| **Time alignment across multi-signal channels** | Alignment based on time stamps of every second |  |  |  |
| **Sliding windows** | Window for signal QC: non-overlapping window for every minute<br>Window for Motion detection: 80% overlap for every five minutes<br>Window for detecting abnormal HR: 80% overlap for every five minutes |  |  |  |

### 5.1 Stream processing module

PAWO ensures efficient and robust data transmissions between the wristband and the smartphone, which is capable of handling large volume of data from diverse sensor types. We made the following modifications to the accompanied SDK/API toolkits: (1) we encoded sampling time as 32-bit Unix timestamp accuracy (achieving the resolution to millisecond), providing sufficient resolution for time alignment across events and among data from different sensor types. (2) We packaged sensor type identifiers with the time stamp as well as the acquired sensor readouts into individual data streams, which are subsequently transmitted to the connected mobile phones. (3) We also made the system automatically check the wearing/idle status of the devices, and this signal is separately transmitted to the connected mobile phones. (4) These signals are packaged and sorted automatically at the backend cloud sever.

### 5.2 Pre-processing module

The pre-processing module was designed for signal quality identification (SQI), denoising, signal smoothing, and adjustment for personal physiological base lines.

#### 5.2.2 Signal Denoising

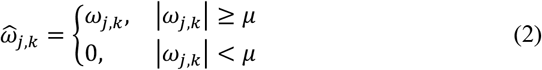

The hard threshold can overcome computing challenges such as singular points in input signals. For seizure detection, singular point signals might contain clinically relevant information. The commonly used VisuShrink method was used to select the threshold.

For signal reconstruction, we adopted the Mallat algorithm to decompose input signals, and designed low-pass and high-pass filters to derive the approximate low-frequency and high-frequency coefficients. After thresholding the high-frequency components, we performed step-wise signal reconstruction as follows:

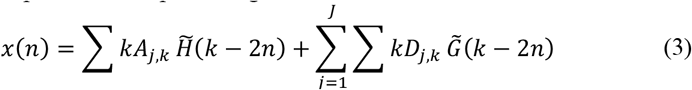

where 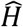 and 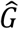 are low-pass filter and high-pass filter respectively, constituting the synthesis filter.

#### 5.2.3 Signal Smoothing

We used moving average to smooth signals and removed extreme data points falling the upper and bottom five percentiles across all the signal spectrum.

#### 5.2.4 Adjusting Personal Physiological Baselines

To account for personal effects on detecting seizure events, we considered personal baseline as the medians of physiological signals from each person, and calculated the residue values by subtracting the medians from the input signals.

### 5.3 Signal analysis module

The signal analysis module has the following components: (1) general analysis to derive overall distributions of the input signals; (2) data analysis for motion detection for three-axis ACC signals; (3) data analysis for detecting abnormal HR/HRV signals.

#### 5.3.1 The general analysis component

Statistical analyses were performed to summarize the overall statistical distribution of signals and variables. These statistic data included maximum, minimum, average, median, standard deviation, Q1 (25% quartile), Q3 (75% quartile), 10% percentile, 90% percentile and peak differences)

#### 5.3.2 Motion detection from ACC signals

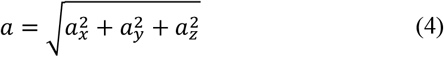

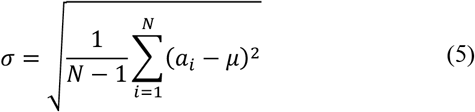

Epochs with σ below 0.1g were considered as resting states, and were removed from further analysis. When five-minute sliding window was used allowing 80% overlapping, there were at most 5 identifiers in an epoch. We took the one with more identifier as the epoch identifier.

#### 5.3.3 HRV analysis

The HR signals were collected for heart rate viability analysis. Epochs with σ below 0.1 g were automatically discarded from further analysis and treated as non-motor state and hence non-seizure events. Next, R-R interval signals (i.e. distance between consecutive heart beats) were calculated from the valid HR signal through the PAWO pipeline. Finally, R-R interval signal in the 5-minute epochs were used to calculate HRV-associated parameters with sliding windows of 5 minutes with 80% overlap (4 minutes). The HRV-associated parameters used in PAWO included the following: standard deviation of normal R-R intervals (SDNN), root mean squared differences of the standard deviation (RMSSD), adjacent normal R-R interval of difference between the standard deviation (SDSD), the number of pairs of successive NNs that differ by more than 50 ms (NN50), the proportion of NN50 divided by total number of NNs (pNN50) in time domain, very low frequency (VLF, 0.001-0.04Hz), low frequency (LF, 0.04-0.15 Hz), high frequency (HF, 0. 15-0.4 Hz), the ratio of low frequency power and high frequency power (LF/HF), total power (TP) in frequency domain. Normalization was conducted on these HRV-associated parameters against the device build-in smoothing algorithm. When 5 minutes sliding window is used with 80% overlapping, there are at most 5 groups of HRV parameters in an epoch. We take the mean value as the groups of HRV parameters.

### 5.4 Multi-signal time alignment module

Using statistical analysis and machine learning to detect seizure onsets is essentially a pattern recognition problem for input signals in the time domain. Analyzing multi-channel signals from different sensor types requires unification of their respective time scale, leveraging the time stamps in our UNIX system. We aligned the signals using sliding windows based on the following resolution:

1. For quality check of input signals, the length of each sliding window was one minute, and there was no overlap between any two windows.
2. For both motion detection and HR/HRV analysis, the length of each sliding window was every five minutes, allowing 80% overlap between adjacent windows.

## 6. Physiome and activity study

The correlation analyses and statistical analyses were conducted on physiological signals to measure physiological changes across childhood developmental stages.

Multiple Linear Regression (MLR) analysis was independently conducted on 4 physiological signals (HR, GSR, ACC magnitude and SDNN) by controlling for age, sex and the time of the day. The pre-processing was performed as follows. First, the time period in which the human subject had a recorded seizure event were excluded from the analysis. Subsequently, the signals were preprocessed in the PAWO pipelines to denoise; outliners were dropped, and data was normalized by established baselines. In each individual MLR model, the response variables were calculated as the mean of the physiological signals in both daytime and nighttime; and the explanatory variables were age, sex and the time of the day. The linear model was written as the following:

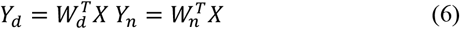

Take HR signal as an example, *Y*_*d*_ ∈ *R*^*n*×1^, *n* = 66 represent the mean HRs of 66 participate in daytime (8 a.m. to 8 p.m.); *Y*_*n*_ ∈ *R*^*n*×1^, *n* = 66 represent the mean HRs in nighttime (9 p.m. to 7.a.m.). Whereas *X* ∈ *R*^*n*×*m*^, *n* = 66, *m* = 3 represented the 3 physiological dimensions (age, sex, the time of the day) of the 66 participants. The **Equation 6** could further aggregate into the following:

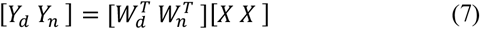

The contribution of age, sex and time were explained via regression coefficients 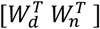; their statistically significances were shown in **Figure 2A**.

Further statistical analyses were conducted on each physiological signal (HR, GSR, ACC and SDNN) to calculate the distribution characteristics across development stages. Boxplots were used to demonstrate the statistical characteristics, and Wilcoxon rank-sum tests were used to estimate statistical significance. The results were shown in **Figure 2** and **Supplemental Figure S6**.

## 7. Principal Component Analysis (PCA) on developmental physiologies during childhood development

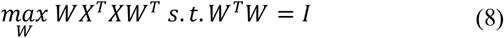

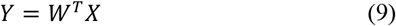

The raw signals from each sensor were processed through PAWO pipeline. SDNN variables and ACC magnitude were also calculated. Then, statistical analyses were conducted on these 7 groups of variables to acquire 66 × 70 (*n* × *m*) size matrix. Next, before implementing PCA algorithm, zero-mean standardization was applied to normalize features into scale [0 − 1]. Finally, since PCA is an unsupervised method, the algorithm would output 66 three-dimensional 66 × 3 (*n* × *d*) samples in **Equation 9** without labeling. The age groups and gender were labeled to highlight the pattern of clustering. The results were shown in **Figure 3**, which indicated clear clustering pattern on age groups while little clustering by gender.

## 8. Construct prediction model for seizure onset

The overall prediction process was shown in **Supplemental Figure S2**. Multiple signals and variables were pre-processed and aligned in PAWO, including 3-axis ACC, 3-axis GYR, HR, GSR, ACC magnitude, GYR magnitude and R-R interval variables. Sliding window technique was applied to condense the data. Personal information and seizure onset times were also added to the window. Statistical procedures and HRV analysis were applied to extract relevant features. A bootstrapping-based ensemble network approach was used to recognize seizure onsets. The problem of seizure detection was posed as a supervised learning task, in which the goal was to classify each 60-second epoch as seizure or non-seizure based on extracted features from EDA and ACM recordings. If any epoch between the start and end of a labeled seizure was correctly classified as a seizure event, the seizure was considered detected (true positive). If multiple epochs within the seizure duration were detected, these were treated as a single true positive. False positives that occurred within 3 minutes from each other were treated as a single false alarm. Details are provided below.

Firstly, sliding window method (60 seconds long) was used to condense the time series into slices. The time window length was set as 60 seconds and overlapping set as 50%, i.e. 30 seconds. The time windows with more than 60% data missing were excluded. Then, according to the seizure events recorded by the nurse or caregivers, the time windows corresponding to seizure onsets were labeled as positive, the remaining windows were labeled as negative. In addition, 60 seconds buffer zones were added to the seizure events; these buffer zones were excluded as negative windows.

Next, features were extracted from each positive or negative window. These features included personal information such as age and sex, time series statistics and HRV variables. The time series parameters included heart rate, R-R interval, 3-axis accelerometer, accelerometer magnitude, 3-axis gyroscope, gyroscope magnitude and galvanic skin response. All the time domain and frequency domain HRV variables were calculated though PAWO. In summary, a total of 120 features from each 60 seconds window was calculated for the subsequent machine learning step to recognize seizure onset. These features are listed as follows:

Lastly, a bootstrapping-based embedding network approach, XGBoost (Chen et al 2016), was adopted from the sci-learn package (Pedregosa et al 2011) and used to overcome the imbalance between negative and positive groups. This model balances the distribution of positive and negative samples by bootstrap sampling and further alleviates the sample bias by embedding independent classifiers. The steps of algorithm are as follows,

The XGboost classifiers in each branch had equal effect. Therefore, the final prediction score of bootstrapping-based ensemble network was averaged over the prediction scores output by all classifiers, i.e.,

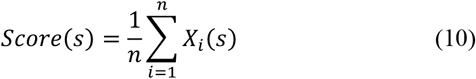

where, *n* is the branch number of ensemble network.

A total of 734,933,434 data points collected from the discovery cohort were used to construct the XGboost model. The dataset was divided into 10-fold for cross-validation; leave-one-seizure-patient-out cross-validation was used. In the whole data set, the patients were divided into ten groups, and the total number of seizure onset was distributed as close as possible among the groups. In each validation process, nine groups were used as the training set, and the remaining one was used as the test set. In the training process, 64 branches were constructed in the bootstrapping-based ensemble network {*X*_*i*_}_*n*=1,2,,..,64_. XGboost classifiers in each branch shared the same parameter settings, which were tuned by the random grid-search technique. The configuration parameters were set as the following: number of estimators were set as 400, learning rate was set as 0.1, scale positive weight was set as 1, silent set was set as 0, subsample set was set as 0.9, regulation coefficients were set as 2 and 0, minimum child weight was set as 3, maximum tree depth was set as 8. The testing process was implemented on the trained XGBoost ensemble network.

Area Under Receiver Operating Characteristic curve (AUROC) was used to evaluate our model. The ROC curve was shown in **Figure 5A**, the contribution of features was shown in **Supplemental Figure S3**.

## 9. Construction of the cloud-based prediction system leveraging the pre-trained model

An alarm system based on cloud computing was implemented using the trained XGBoost ensemble network with the optimized decision-making threshold. The alarm system possessed the complete training ensemble network {*S*_*i*_}_*n*=1,2,,..,64_ from the blind validation procedure. All the samples were subsequently tested on our real-time data. The real-time data steam was continuously transmitted from wearable sensors to the backend cloud. In the cloud sever, PAWO executed the prediction algorithm every thirty second to analyze input signal at a time interval of 60 seconds. With the pre-trained model, we can immediately compute the prediction scores. When prediction scores were beyond our optimized threshold, the system immediately sent out warning messages to caregivers. Fast data transmission, cloud synchronization and automatic prediction has enabled fast decision making almost right after any abnormal physiological signals captured by the machine.

The alarm system was applied on 33 independently recruited children (whom were not involved in training the model). A total of 297,651,517 wearable sensor data points from these individuals were screened by our cloud system, and 11 out of the 33 children were recorded at least one seizure event. We compared the systems warning signal against clinical observations as well as against EEG signals whenever possible.

